# Gut microbiota modulates distal symmetric polyneuropathy in diabetic patients

**DOI:** 10.1101/2022.10.07.22280806

**Authors:** Junpeng Yang, Xueli Yang, Guojun Wu, Fenglian Huang, Xiaoyang Shi, Wei Wei, Yingchao Zhang, Haihui Zhang, Lina Cheng, Lu Yu, Jing Shang, Yinghua Lv, Xiaobing Wang, Rui Zhai, Pan Li, Bota Cui, Yuanyuan Fang, Xinru Deng, Shasha Tang, Limin Wang, Qian Yuan, Liping Zhao, Faming Zhang, Chenhong Zhang, Huijuan Yuan

## Abstract

The contribution of the gut microbiota to distal symmetric polyneuropathy (DSPN) in diabetic patients remains elusive. We found that the gut microbiota from DSPN patients induced more severe peripheral neuropathy in db/db mice. Gut microbiota from healthy donors significantly alleviated DSPN independent from glycaemic control in patients in a randomized, double-blind and placebo-controlled trial. The gut bacterial genomes correlated with Toronto Clinical Scoring System score were organized in two competing guilds. Increased Guild 1 that had higher capacity in butyrate production and decreased Guild 2 that harbored more genes in synthetic pathway of endotoxin were associated with improved gut barrier integrity and decreased proinflammatory cytokine levels. Moreover, matched enterotype between transplants and recipients showed better therapeutic efficacy with more enriched Guild 1 and suppressed Guild 2. Thus, the two competing guilds may mediate the causative role of the gut microbiota in DSPN and have the potential to be an effective target for treatment.

## INTRODUCTION

As the most common neuropathy in patients with diabetes mellitus (DM), distal symmetric polyneuropathy (DSPN) is a pernicious unmet medical need, which affects approximately half of the diabetic patients including both type1 and type 2 diabetes mellitus (T1DM and T2DM) (Hicks and Selvin, 2019; Sloan et al., 2021). DSPN is associated with increased mortality, lower-limb amputations and distressing painful neuropathic symptoms, which negatively affect patients’ functionality, mood, and health-related quality of life (Li et al., 2013; Slangen et al., 2014). Although lifestyle modification, glucose control and a few medications may ameliorate the symptoms in some patients with DSPN, no disease-modifying treatments for DSPN are available due to the paucity of understanding of its underlying pathogenetic mechanism and lack of effective targets (Pop-Busui et al., 2017; Sloan et al., 2021).

Compelling and converging evidence show a causative role of gut microbiota in maintaining the glucose homeostasis (de Groot et al., 2020; Metwaly et al., 2022; Sanna et al., 2019; Tang et al., 2022; Zhao et al., 2018). The gut microbiota may also function at the intersection between the gut-brain and the neuroimmune-endocrine axes, forming a complex network that can affect the nervous system (Fung et al., 2017; Pane et al., 2022). Increasingly, studies have demonstrated that the cross-talk between the gut microbiota and brain may crucially impact neurodegenerative disorders of the central nervous system, such as in Parkinson’s and Alzheimer’s diseases (Bulgart et al., 2020; Tansey et al., 2022). In addition, the gut microbiota and its bioactive substances are known to regulate host bioenergetics and inflammation, possible mechanistic pathways of DSPN (Bonhof et al., 2019; Thevaranjan et al., 2017; Tilg et al., 2020). Recently, a cohort study with small sample size showed that the gut microbiota of patients with DSPN was different from healthy control (Wang et al., 2020). However, the causative relationship between the gut microbiota and peripheral nervous system disorders in DM remains elusive.

Fecal microbiota transplantation (FMT) from healthy donors to the gut of patients and/or from patients to mice has become a powerful strategy to demonstrate if the gut microbiota may play a causative role in a particular pathology such as DSPN (Hanssen et al., 2021; Sorboni et al., 2022). Here, we found significant differences of the gut microbiota among the subjects with normal glucose level and DM patients with or without DSPN. Furthermore, we showed that the fecal microbiota from patients with DSPN induced more serious peripheral neuropathy than the subjects with normal glucose level by inoculating the fecal microbiota to the db/db mice, suggesting that dysbiosis of gut microbiota contributes to the progression of DSPN. Then we performed FMT from healthy donors to DSPN patients who responded poorly to conventional treatments in a randomized, double-blind, placebo-controlled pilot clinical trial. The results showed that modulation of the composition and function of gut microbiota via FMT from healthy donors induced significant alleviation of DSPN. This study revealed a potentially causative link between the gut microbiota and peripheral nervous system disorders in DM, which could become a therapeutic target for developing effective treatment.

## RESULTS

### The gut microbiota from patients with DSPN induced more severe peripheral neuropathy in db/db mice

We collected fecal samples from the DM patients with DSPN (DSPN, n = 27), and compared their gut microbiota with that in DM patients without DSPN (DM, n = 30) and subjects with normal glucose level (NG, n = 29) from our previous dataset (Deng et al., 2022; Fang et al., 2021) (Table S1). Based on the sequencing data of V3-V4 regions of 16S rRNA genes, the richness of gut microbiota in patients with DSPN was significantly higher than that in DM and NG groups (Figure S1A). Principal coordinate analysis (PCoA) based on Jaccard distance showed that the overall structure of gut microbiota in DSPN was different from that in DM and NG groups (Figure S1B).

Next, to evaluate whether the dysbiosis of gut microbiota contributes to the development of peripheral neuropathy, we transplanted the fecal microbiota from DSPN and NG group to the genetically diabetic mice respectively. After treatment with a cocktail of antibiotics for 2 weeks, the db/db mice at 13-weeks old were gavaged with the fecal microbiota either from DSPN (M-DSPN, n = 6) or NG group (M-NG, n = 6). The structure of the gut microbiota in the M-DSPN mice was significantly different from that in M-NG mice (*P* = 0.004 by PerMANOVA test with 999 permutations), and the gut microbiota of the recipient mice was more similar to that of the corresponding human donors (Figure S2). We evaluated the severity of peripheral neuropathy in the mice at the end of third week after FMT. Compared to the M-NG group, the M-DSPN mice showed higher mechanical and thermal sensitivity, which were reflected by the level of 50% threshold and thermal latency, and lower MNCV of sciatic nerve (Figure 1A). The M-DSPN mice also had significant lower intraepidermal nerve fiber density (IENFD) of the posterior plantar skin by immunohistochemistry analysis, which was used to assess the neuropathy in the small fiber (Figure 1B). In the peripheral nervous systems, neurofilament 200 (NF200) was a special marker for A-fibers, myelin basic protein (MBP) was associated with the formation and maintenance of myelin, and brain derived neurotrophic factor (BDNF) was required for survival, proliferation, migration and differentiation of neurons and neurogliocytes (Fan et al., 2020; Zhang et al., 2019) (McGregor and English, 2018). Previous studies have found that NF200, MBP and BDNF decreased significantly in dorsal root ganglion (DRG) and sciatic nerve when peripheral nerve injured by diabetes (Fan et al., 2020; Zhang et al., 2019).The levels of these three proteins were significantly lower in M-DSPN mice than M-NG group both in DRG and sciatic nerve (Figure 1C and D). The above results suggest that the gut microbiota from patients with DSPN aggravates peripheral neuropathy in db/db mice.

**Figure 1.**
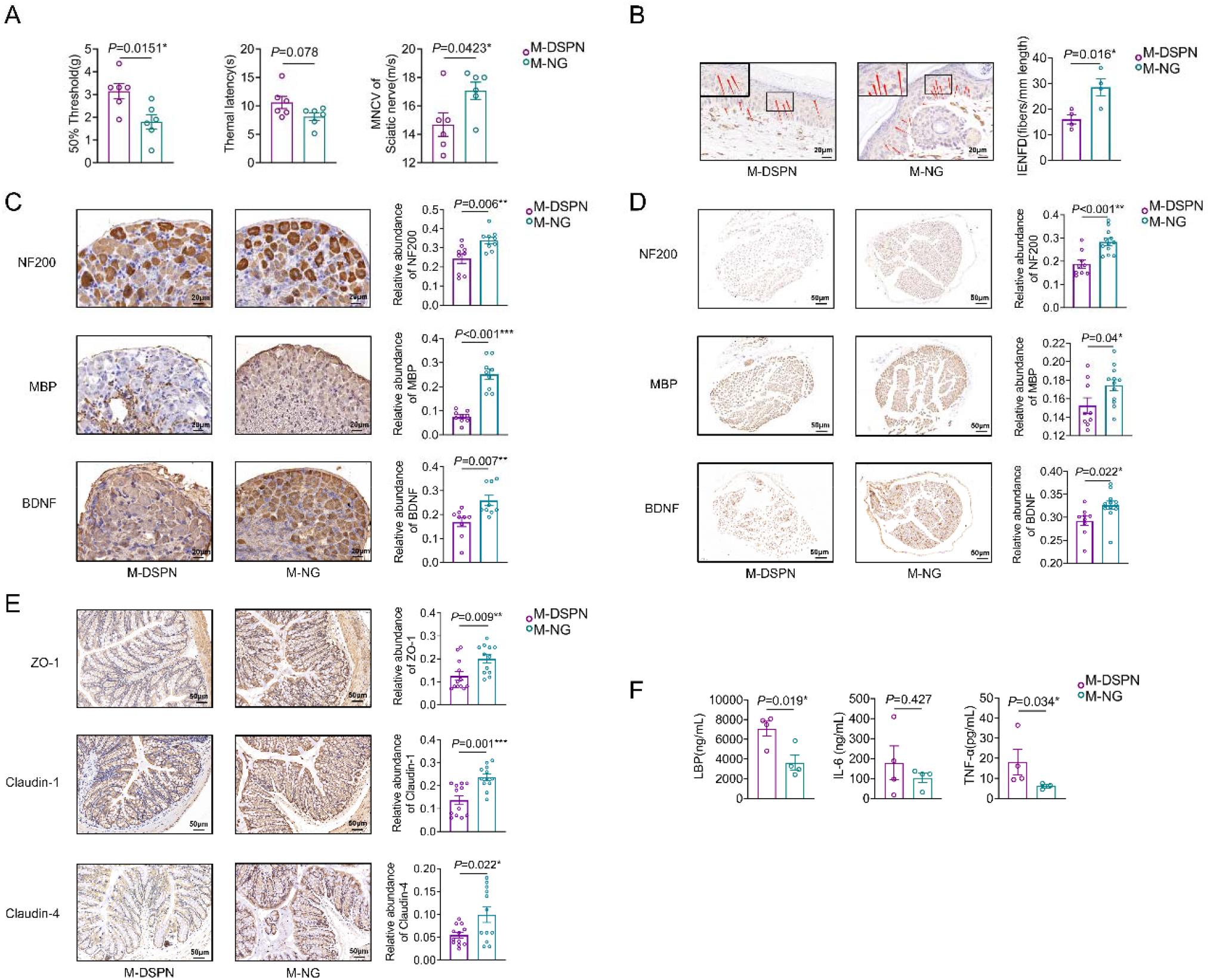
Gut microbiota from patients with DSPN induced more severe peripheral neuropathy than that from the subjects with normal glucose level in db/db mice. **(A)** Mechanical sensitivity, thermal sensitivity and motor nerve conduction velocity (MNCV) of sciatic nerve. Immunohistochemical staining of the posterior plantar skin and integrated optical density (IOD) analysis for **(B)** Intraepidermal nerve fibre density (IENFD, marked by PGP9.5). Immunohistochemical staining of **(C)** dorsal root ganglion and **(D)** sciatic nerve and IOD analysis for Neurofilament 200 (NF200), myelin basic protein (MBP) and Brain derived neurotrophic factor (BDNF). **(E)** Immunohistochemical staining of clone and IOD analysis for tight junction proteins ZO-1, Claudin-1 and Claudin-4. **(F)** Plasma levels of LBP, IL-6 and TNF-α. Data are presented as the mean ± s.e.m. The comparison between the two groups was tested by Student’s t-test (two-tailed). * *P* < 0.05, ** *P* < 0.01 and *** *P* < 0.001. M-DSPN indicates db/db mice received fecal microbiota from patients with DSPN, M-NG indicates db/db mice received fecal microbiota from the subjects with normal glucose level. For A, M-DSPN group, n = 6; M-NG group, n = 6; For B and F, M-DSPN group, n = 4; M-NG group, n = 4; For C, M-DSPN group, n = 3; M-NG group, n = 3; For D and E, M-DSPN group, n = 3; M-NG group, n = 4. For B and C, scale bars indicate 20 μm; For D and F, scale bars indicate 50 μm.

We also assessed the effects of gut microbiota from patients with DSPN on the gut barrier function. The immunofluorescence staining showed that the expression of the tight junction proteins ZO-1, Claudin-1 and Claudin-4 in the colonic mucosal biopsy specimens were significantly lower in M-DSPN mice than that in M-NG group (Figure 1E), indicating worse gut barrier function in M-DSPN mice. Then we tested the serum level of lipopolysaccharide (LPS)-binding protein (LBP), which can bind to antigens such as endotoxin produced by bacteria and represent a surrogate biomarker that links bacterial antigen load in the blood and the host inflammatory response. Notably, the level of LBP was significantly higher in the M-DSPN mice than that in M-NG group (Figure 1F). We assessed the inflammatory status of the mice. Indeed, the serum levels of TNF-α and IL-6, which are systemic inflammation biomarkers that have been associated with DSPN progression (Feldman et al., 2017; Herder et al., 2017), were higher in the M-DSPN mice than that in M-NG group (Figure 1F). These results suggest that gut barrier dysfunction, higher antigen load and severer systemic inflammation may contribute to the aggravation of peripheral neuropathy by the gut microbiota from patients with DSPN.

### FMT alleviated the severity of peripheral neuropathy in patients with DSPN

As the dysbiosis of gut microbiota contributed to the development of peripheral neuropathy, we would like to find out whether improvement of gut microbiota can alleviate DSPN in patients. Thirty-seven DSPN patients were randomized into either FMT or placebo group in a 2:1 ratio. Before enrolment into this trial, all patients showed no improvement in their DSPN symptoms after at least 84 days of conventional treatments (lifestyle modification, glucose control and drug intervention, medical details in Table S2). After the run-in period, these patients were transplanted with transplants made from fecal microbiota of the healthy donors or placebo and then were followed up for 84 days. Finally, 22 patients in the FMT group and 10 patients in the placebo group completed this RCT study (Figure S3). At baseline (0D), demographic and anthropometric characteristics and glycemic control were similar between the two groups (Table S3). In particular, there were no significant differences in the level of Toronto Clinical Scoring System (TCSS (Arumugam et al., 2016)) score and the sensory and motor nerve functional status of DSPN (Figure 2 and Figure S4). At 84 days after transplantation (84D), glucose and lipid metabolism, blood pressure and body weight remained unchanged in both groups (Table S3). The insulin requirement was similar between the two groups (Table S3) and no serious adverse events were documented during the trial (Table S4).

**Figure 2.**
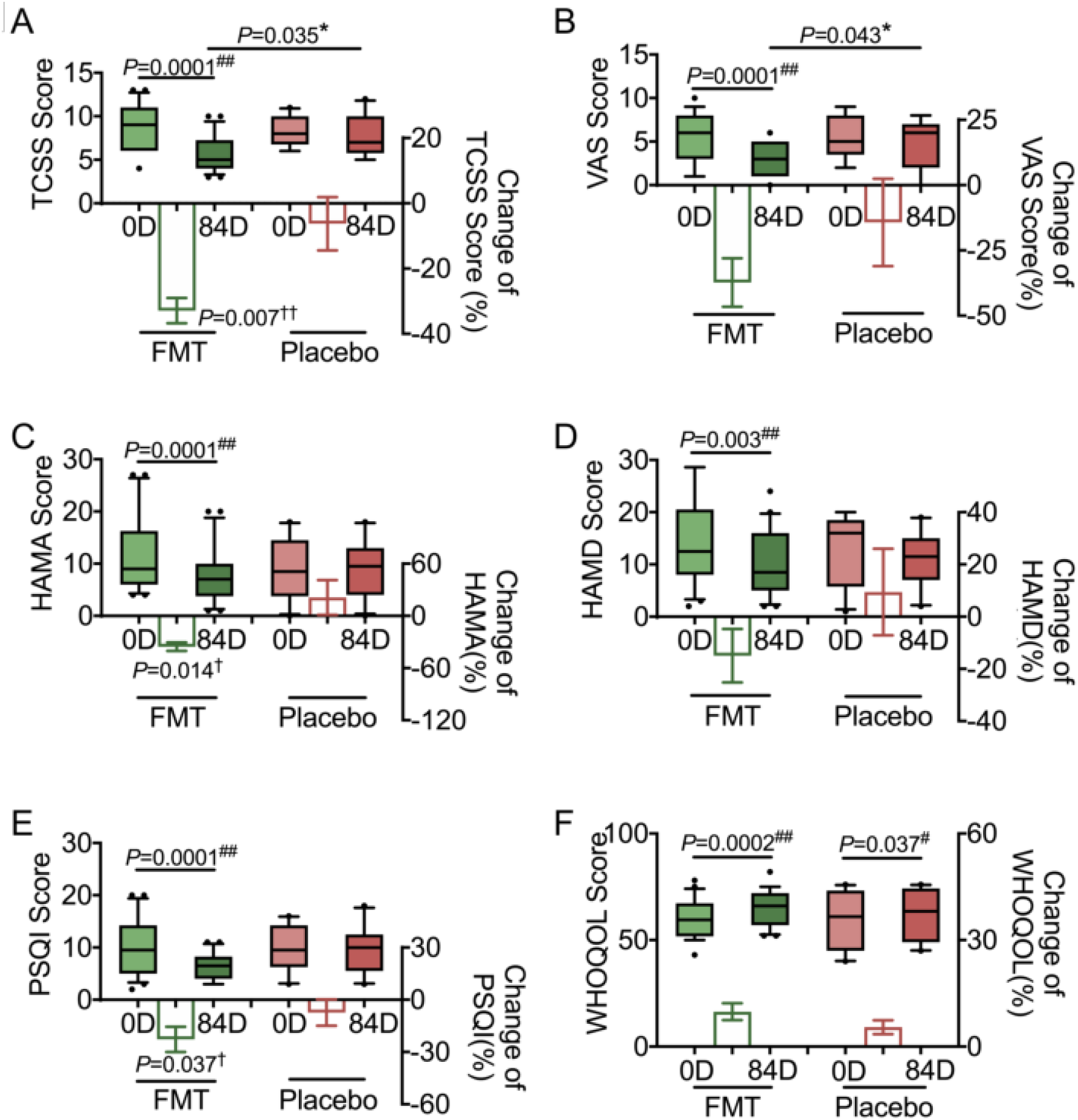
FMT alleviated the severity of peripheral neuropathy, anxiety, depression, and improved sleep and life quality in patients with DSPN. **(A)** Toronto clinical scoring system (TCSS) score. **(B)** Visual analogue scale (VAS) score. **(C)** Hamilton anxiety scale (HAMA) score; **(D)** Hamilton depression rating scale (HAMD) score. (**E**) Pittsburgh sleep quality index (PSQI) score. (**F**) World health organization’s quality of life (WHOQOL)-BREF score. 0D indicates baseline, and 84D indicates 84 days after FMT. In the box plots, the line in the middle of the box is plotted at the median, and the inferior and superior limits of the box correspond to the 25^th^ and 75^th^ percentiles, respectively. The whiskers correspond to the 10^th^ and 90^th^ percentiles, and outliers are denoted. Mann-Whitney U test was used to analyze differences between FMT and Placebo. * *P* <0.05. Wilcoxon matched-pairs signed rank test was used to analyze each pairwise comparison 0D and 84D within each group. ^#^ *P*<0.05 and ^##^ *P*<0.01. The bars represent the mean change from the baseline value per group, with the corresponding s.e.m.. Mann-Whitney U test was used to analyze differences in the changes between the FMT and placebo groups (intergroup changes). ^†^*P* < 0.05 and ^††^*P* < 0.01. FMT group, n = 22; Placebo group, n = 10.

TCSS score is used to evaluate the neuropathic symptoms and signs of patients, as the standard to assess the severity of DSPN (Arumugam et al., 2016). We used the change of TCSS score at 84D compared with baseline as the primary outcome and the results showed that the reduction of TCSS score was significantly greater in the FMT group than that in the placebo group (Figure 2A). Moreover, the levels of TCSS at 84D for the patients subjected to FMT were significantly lower than baseline, but no changes were observed in the placebo group, and the levels of TCSS in the FMT group were significantly lower than that in the placebo group at 84D (Figure 2A). We also evaluated neuropathic pain of the patients with visual analogue scale (VAS (Petersen et al., 2021)) score. Compared to baseline, the VAS scores also decreased significantly after FMT, but not in placebo group. The levels of VAS were significantly lower than that in the placebo group at 84D (Figure 2B). Notably, fifteen patients in the FMT group, who suffered from moderate/severe neuropathic pain (VAS score ⩾4) at baseline and eight of them (53.3%) had more than 50% pain relief at 84D as compared with baseline, which was considered a good outcome (Petersen et al., 2021). While only 14.29% patients in placebo group showed more than 50% pain relief at 84D, based on the VAS, which was significantly lower than that in FMT group. These results indicate that FMT induced stable relief of the symptoms and signs, particularly the neuropathic pain, in the patients with DSPN.

As the neuropathic symptoms severely induce anxiety, depression, sleep disorders and reduce the quality of life in DSPN patients (Gylfadottir et al., 2020), we assessed the effects of FMT on anxiety, depression, sleep quality and overall quality of life by using the Hamilton Anxiety Scale (HAMA (Zhao et al., 2021)), Hamilton Depression Rating Scale (HAMD (Zhao et al., 2021)), Pittsburgh Sleep Quality Index (PSQI (Buysse et al., 1989)) and the Brief table of the World Health Organization’s Quality of Life (WHOQOL-BREF (Saxena et al., 2001)). After 84 days of transplantation, HAMA and HAMD score showed that the anxiety and depression status were significantly improved in the FMT group but not in the placebo group (Figure 2C and D). PSQI assessment showed that sleep quality was significantly improved in patients subjected to FMT at 84D but not in those that received the placebo (Figure 2E). The WHOQOL-BREF score significantly increased in both FMT and placebo groups at 84D, suggesting improved quality of life for all the patients (Figure 2F).

We used electrophysiological measurements (nerve conduction velocity (NCV) and current perception threshold (CPT)) to objectively assess whether FMT improve peripheral neuropathy (Dyck et al., 1997). At 84 days after transplantation, the sensory nerve conduction velocities (SNCVs) of sural nerves and ulnar nerves were significantly increased only in patients of the FMT group and the changes of SNCVs were significantly greater in the FMT group than in the placebo group (Figure S4A-C). The motor nerve conduction velocities (MNCVs) of the distal median nerves, posterior tibial nerves and common peroneal nerve were significantly improved in patients of the FMT group but not in placebo (Figure S4D-G). CPT can be used to visualize the sensitivity of different types of never fibers to electrical stimulation, including the thick myelinated (Aβ) fibers (2000Hz), thin myelinated (Aδ) fibers (250Hz), and unmyelinated (C) fibers (5Hz) (Lv et al., 2015). For superficial and deep peroneal nerve, the CPT level at 5 Hz was significantly decreased in FMT group after 84 days, and significantly lower in FMT group than that in placebo group, indicating that the hypoesthesia of C-fibers in legs of FMT group improved (Figure S4H-J). For distal median nerve, the CPT levels at 2000Hz, 250Hz and 5Hz all decreased significantly in FMT group but not in placebo after 84 days, and the changes at 250Hz and 5Hz were significantly greater in the FMT group than in the placebo group, suggesting that FMT may influence on Aβ, Aδ and C nerve fibers of distal upper limbs (Figure S4K-M). This suggests that introduction of gut microbiota from healthy donors improved the electrophysiological functions of peripheral nerves in patients with DSPN.

After finishing the RCT study, to further verify the alleviation of neuropathic symptoms and signs in patients with DSPN by FMT, we transplanted the transplants made from fecal microbiota of the healthy donors to all of the ten patients in placebo group (post-RCT study). Compared to the baseline or the end of the RCT study, these patients showed significantly improvement of the neuropathic symptoms and signs, anxiety status, sleep quality, overall quality of life, and electrophysiological functions of peripheral nerves after 84 days of transplantation with the transplants from healthy donors (Figure S5 and S6).

### FMT improved gut barrier integrity and systemic inflammatory status in patients with DSPN

As we had found that the dysbiosis gut microbiota from the patients with DSPN made damage of gut barrier in the aforementioned db/db mice study, in order to find out whether FMT, the gut microbiota – targeted treatment, could improve gut barrier function and decrease chronic inflammation of the patients, we collected biopsy specimens of colon from part of our patients at baseline and 84 days after transplantation in RCT study. The immunofluorescence staining showed that the expression of the tight junction proteins ZO-1 and Claudin-1 in the colonic mucosa of biopsy specimens were significantly higher in FMT than that in placebo group at 84D (Figure 3A and B). In addition, Claudin-4 was promoted in specimens from patients in both FMT and placebo group, but the changes were significantly greater in the FMT group (Figure 3C).

**Figure 3.**
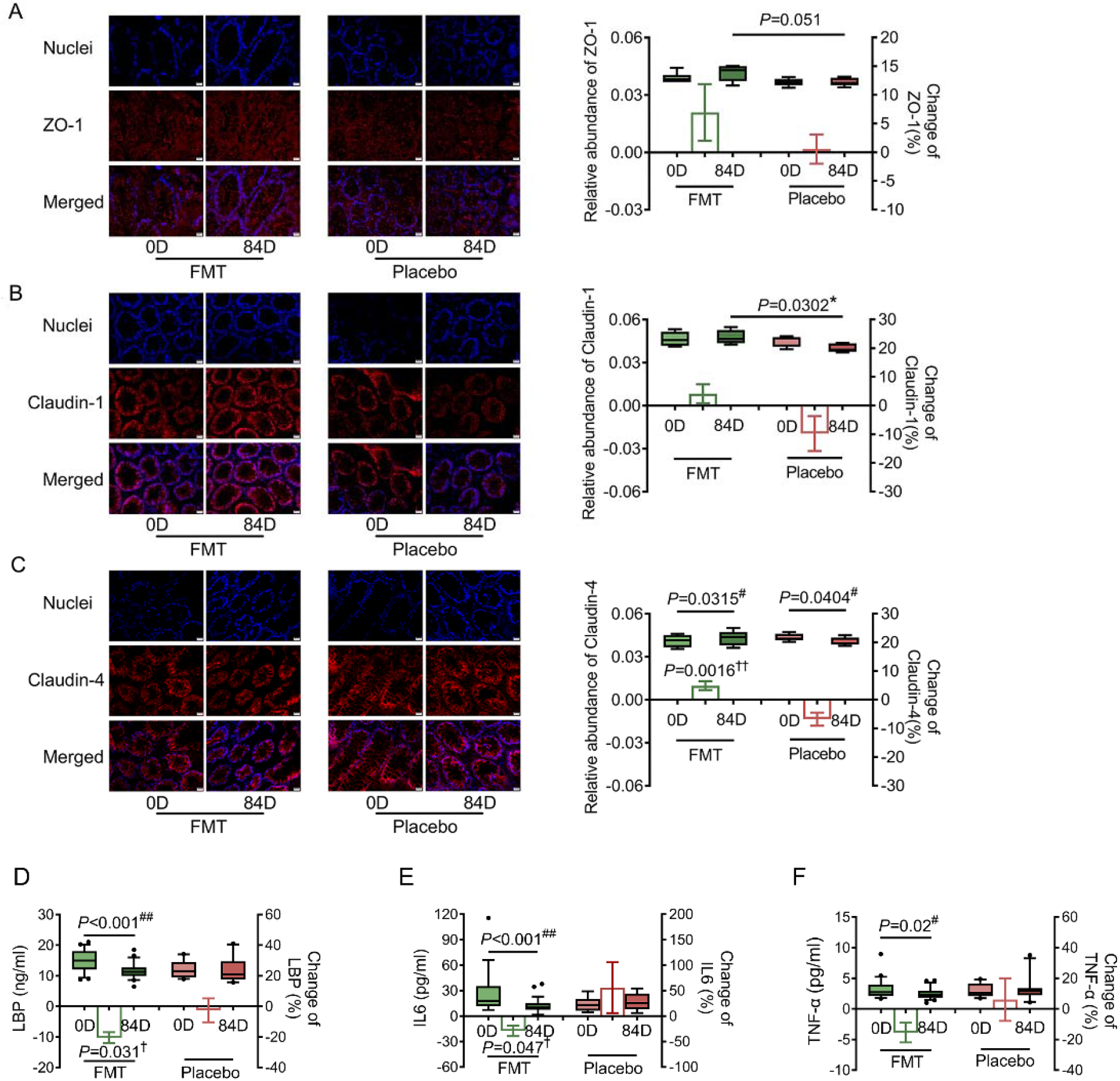
FMT improved gut barrier integrity and systemic inflammatory status in patients with DSPN. Representative immunofluorescence staining of the tight junction proteins **(A)** ZO-1, **(B)** Claudin-1 and **(C)** Claudin-4 in clone. Plasma levels of **(D)** LBP, **(E)** IL-6 and **(F)** TNF-α. Scale bars indicate 20 μm. 0D indicates baseline, 84D indicates 84 days after FMT. In the box plots, the line in the middle of the box is plotted at the median, and the inferior and superior limits of the box correspond to the 25^th^ and 75^th^ percentiles, respectively. The whiskers correspond to the 10^th^ and 90^th^ percentiles, and outliers are denoted. Student’s t-test (two-tailed) (A, B and C) and Mann-Whitney U test (D, E and F) were used to analyze differences between FMT and Placebo. * *P* < 0.05. Paired t-test (two-tailed) (A, B and C) and Wilcoxon matched-pairs signed rank test (D, E and F) was used to analyze each pairwise comparison 0D and 84D within each group. ^#^ *P* < 0.05 and ^##^ *P* < 0.01. The bars represent the mean change from baseline per group, with the corresponding s.e.m. Student’s t-test (two-tailed) (A, B and C) and Mann-Whitney U test (D, E and F) were used to analyze differences in the changes between the FMT and placebo groups (intergroup changes). ^†^ *P* < 0.05 and ^††^ *P* < 0.01. For A and C, FMT group, n = 6; placebo group, n = 5. For B, FMT group, n = 4; placebo group, n = 5. For D, E and F, FMT group, n = 22; placebo group, n = 10.

Because the improvement of the gut barrier integrity would decrease the serum antigen load from microbiota, we tested the serum level of LBP, and found that the level of LBP was significantly reduced in the FMT group but not in the placebo group (Figure 3D). Then we assessed the inflammatory status of patients with DSPN after FMT. Indeed, the serum levels of IL-6 and TNF-α decreased in the FMT group at 84D but did not change in the placebo group (Figure 3E and F). This suggests that FMT improved the gut barrier function, reduced serum antigen load and aforementioned alleviation of chronic inflammation, which may contribute to alleviate DSPN in patients.

### FMT induced overall structural changes in the gut microbiota in patients with DSPN

To further explore the role of gut microbiota in the beneficial effect on DSPN alleviation via FMT, we did shotgun metagenomic sequencing of all the transplants made from gut microbiota of the donors, and the fecal samples from all the patients in the RCT study and post-RCT study before transplantation (0D), 3 days (3D), 28 days (28D), 56 days (56D), and 84 days (84D) after transplantation (Table S5). To characterize the strain-level changes in the gut microbiota in patients following FMT, we *de novo* assembled 1,572 high-quality draft genomes from the metagenomic dataset (details in Materials and Methods, Table S6 and S7). After integrating the genomes from Human Gastrointestinal Bacteria Genome Collection (HGG) (Forster et al., 2019) to improve the metagenomic analysis, 1,999 non-redundant high-quality metagenome-assembled genomes (HQMAGs) were obtained and used in the further analysis (Table S8). Transplantation with placebo showed no significant alteration in the gut microbiota in the patients during the RCT study (Figure S7). Then samples from the FMT group in RCT study and sample from the post-RCT study together with transplants from healthy donors were analyzed to reveal the alteration in gut microbiota during FMT. To determine probable microbial origins in longitudinal samples after FMT, unique HQMAGs detectable in pre-FMT patient (0D) samples and FMT transplants were compared (Figure 4A). The relative abundance of HQMAGs only detected in the FMT transplants only accounted for 48.1% of the HQMAGs shared between the 0D samples of patients and corresponding transplants. Transplant-derived HQMAGs accounted for a considerable proportion of the population from day 3 to day 84 after FMT (3D to 84D). The distance between the patient’s gut microbiota and that of corresponding transplants was significantly decreased after FMT, although the gut microbiota structures in the patients were still closer to each individual’s own baseline composition than to the transplants (Figure 4B). There was a significant increase in richness of the gut microbiota at 28D after FMT, but the diversity measured by Shannon index was not altered (Figure 4C). Subject adjusted principal coordinate analysis (aPCoA) of Jaccard distances at the HQMAGs level demonstrated that the gut microbiome of patients significantly shifted after 3 days of FMT (3D). The gut microbiome further significantly changed after 28 days (28D) and remained relative stable afterward (56D-84D) (Figure 4D). These results suggest that FMT induced early and significant alterations in the gut microbiota of DSPN patients, which were concomitant with significant and stable improvement of neuropathic symptoms and signs.

**Figure 4.**
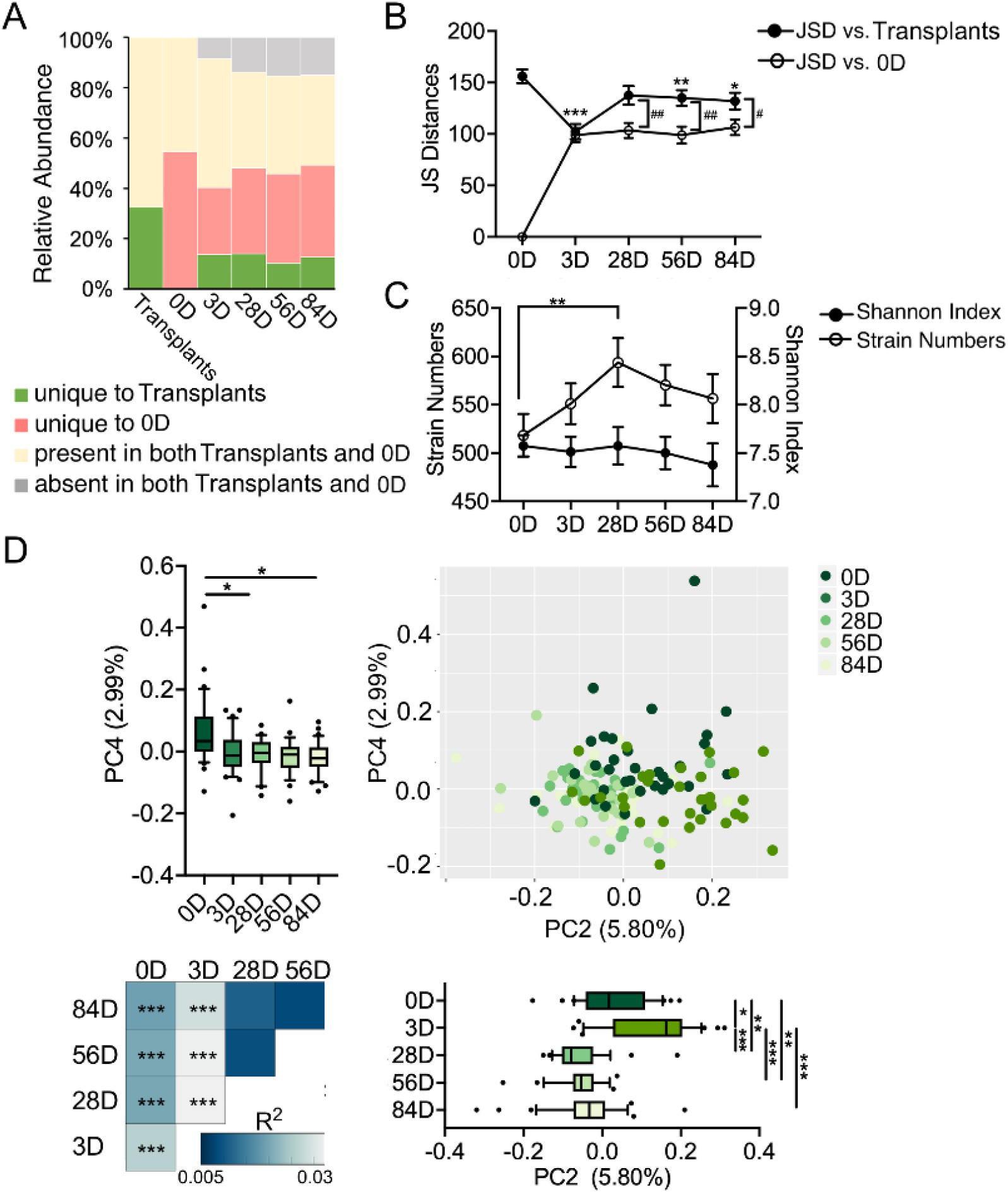
FMT induced overall structural changes in the gut microbiota in patients with DSPN. **(A)** Strains from samples were classified by origin (unique to patient at baseline, unique to transplant, present in both patient baseline and transplant, or absent in both patient baseline and transplant). **(B)** Jensen-Shannon distance (JSD) of the gut microbiotas. JSD vs. donor: JSD between a sample at the particular time point and that of the transplant; JSD vs. T0: JSD between a sample at the particular time point and the baseline sample. Data are presented as the mean ± s.e.m. Wilcoxon matched -pair signed-rank tests were used to analyze each pairwise comparison. \**P* < 0.05 and \*\**P* < 0.01. **(C)** Richness (number of strains) and diversity (Shannon index) of the gut microbiota. Data are presented as the mean ± s.e.m. Wilcoxon matched-pair signed-rank tests were used to analyze each pairwise comparison. \**P* < 0.05 and \*\**P* < 0.01. **(D)** Subject adjusted principal coordinate analysis (aPCoA) of Jaccard distances at the strain level. The lower triangular heat map shows the marginal PerMANOVA test based on Jaccard distance (strata in subject, 999 permutations, \**P* < 0.05 and \*\**P* < 0.01.). The box plots show the changes in the gut microbiota from different time points on PC1 or PC2 (the line in the middle of the box is plotted at the median, the inferior and superior limits of the box correspond to the 25th and 75th percentiles, the whiskers correspond to the 10^th^ and 90^th^ percentiles, and outliers are denoted). Wilcoxon matched-pair signed-rank tests were used to analyze each pairwise comparison. \**P* < 0.05, \*\**P* < 0.01 and \*\*\**P* < 0.001. The samples from FMT group in RCT study and samples from Placebo group in the post-RCT study of transplantation with transplants from healthy donors were analyzed together. 0D indicates before transplantation (n = 32), 3D (n = 31), 28D (n = 29), 56D (n = 26) and 84D (n = 32) indicate 3 days, 28 days, 56 days and 84 days after transplantation with transplants from healthy donors, respectively.

### The gut microbial genomes correlated with TCSS score were organized in two competing guilds

Next, we identified that TCSS scores were significantly correlated with 54 HQMAGs (Figure 5A and Table S9), among which 21 and 33 HQMAGs had negative and positive correlations, respectively, by using linear mixed effect model via MaAsLin2 (Mallick et al., 2021). Most of the HQMAGs (19 of 21) negatively correlated with TCSS score were belonged to Firmicutes, and many of them are potentially beneficial bacteria that can regulate mucosal barrier function to reduce gut permeability and/or modulate the host immune response, possibly via production of butyrate (Cani et al., 2022; Derrien et al., 2022; Gomes et al., 2018; Li et al., 2019; Maioli et al., 2021; Waters and Ley, 2019), such as *Fecalibacterium prausnitzii, Agathobacter rectalis, Agathobaculum butyriciproducens* and *Anaerobutyricum hallii*. Out of the 33 HQMAGs positively correlated with TCSS score, 26 were belonged to Bacteroidetes, in especial 17 from *Bacteroides uniformis*. As bacteria in the gut ecosystem interact with each other and work as coherent functional groups (a.k.a “guilds”) (Wu et al., 2021), we applied co-abundance analysis on these 54 HQMAGs to explore the interactions between them and to find potential guilds structure. The 54 HQMAGs were organized into two guilds-the 21 HQMAGs negatively correlated with TCSS score were positively interconnected with each other and formed as Guild 1, which was true for the 33 HQMAGs positively correlated with TCSS score as Guild 2 (Figure 5B). There were only negative correlations between the two guilds, suggesting a potentially competitive relationship between the two guilds. We found that the abundance of Guild 1 was almost equal to Guild 2 in the gut microbiota of transplants from healthy donors, but the abundance of Guild 2 was significantly higher than Guild 1 in the patients with DSPN at baseline (Figure 5C). Compared to their transplants, the gut microbiota of patients with DSPN contained significantly much more Guild 2 and slightly fewer Guild 1 (Figure 5C). Notably, FMT significantly decreased the abundance of Guild 2 and mildly increased the abundance of Guild 1 from 3D, and there was no significant difference between Guild 1 and 2 from 28D to 84D (Figure 5C).

**Figure 5.**
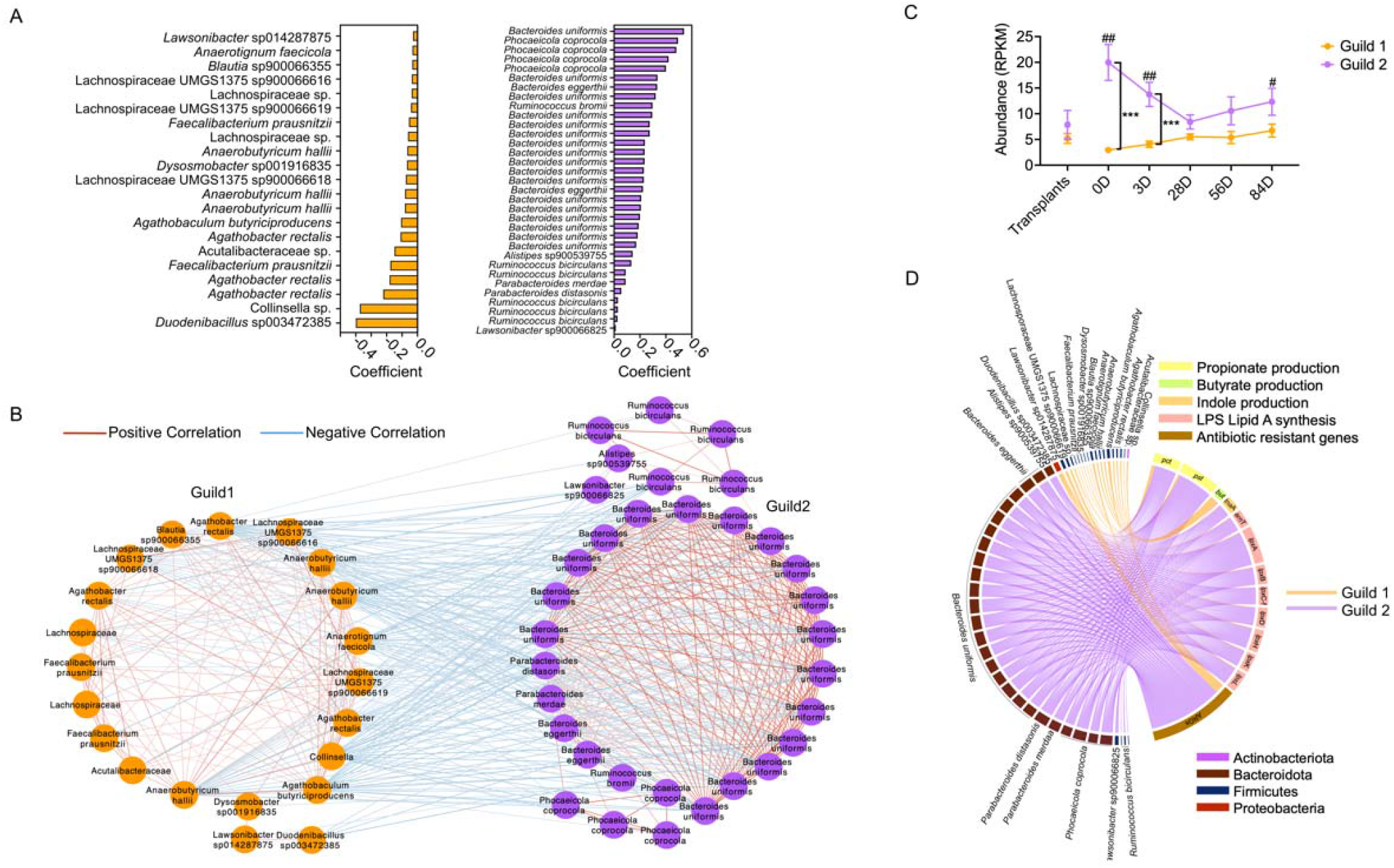
The gut microbial genomes correlated with TCSS score were organized in two competing guilds. **(A)** The high-quality metagenome-assembled genomes (HQMAGs) had significantly correlation with TCSS score. The orange bar indicates negative correlation and purple bar indicates positive correlation. **(B)** Co-abundance network of the HQMAGs reflects two competing guilds. The co-abundance correlation between the HQMAGs were calculated using repeated measures correlation. All significant correlations with BH-adjusted *P* < 0.05 were included. Edges between nodes represent correlations. Red and blue colors indicate positive and negative correlations, respectively. Node color indicates Guild 1 (orange) and Guild 2 (purple), respectively. **(C)** The abundance of Guild 1 and Guild 2 in transplants and fecal samples from patients. The samples from FMT group in RCT study and samples from Placebo group in the post-RCT study of transplantation with transplants from healthy donors were analyzed together. 0D indicates before transplantation (n = 32), 3D (n = 31), 28D (n = 29), 56D (n = 26) and 84D (n = 32) indicate 3 days, 28 days, 56 days, and 84 days after transplantation with transplants from healthy donors, respectively. Data are presented as the mean ± s.e.m. Wilcoxon matched-pair signed-rank tests (two-sided) were used to compare differences of abundance between Guild 1 and 2 at the same timepoint, \*\*\**P* < 0.001. Mann-Whitney tests were used to compare transplants and recipients, ^*#*^ *P* < 0.05 and ^#*#*^ *P* < 0.01. **(D)** Functional genes in the HQMAGs significantly correlated with TCSS score. The encode key enzymes in the propionate, butyrate and indole production, lipid A biosynthesis pathways and antibiotic resistant genes are shown in the right part of the cycle. The HQMAGs that were significantly corelated with TCSS score and whose genomes contain these genes are shown in the left part of the cycle. The linkage between strains and genes indicates that the genome of the strain contained the gene. The orange line indicates that the HQMAGs negatively corelated with TCSS score, whereas the purple line indicates that the HQMAGs positively corelated with TCSS score pst: propionyl-CoA:succinate-CoA transferase; pct: propionate CoA transferase; but: butyryl–coenzyme A (butyryl-CoA):acetate CoA transferase; tnaA: tryptophanase; arnT: 4-amino-4-deoxy-L-arabinose transferase; lpxA: UDP-N-acetylglucosamine acyltransferase; lpxB: lipid A disaccharide synthase; lpxCf (lpxC-fabZ): bifunctional enzyme LpxC/FabZ; lpxD: UDP-3-O-(3-hydroxymyristoyl)glucosamine N-acyltransferase; lpxH: UDP-2,3-diacylglucosamine hydrolase; lpxK: tetraacyldisaccharide 4’-kinase; lpxL: lipid A biosynthesis lauroyltransferase; ARGs: antibiotic resistant genes.

Then we studied the genomic features of the two guilds to understand the potential functions underlying their associations with the alleviation of DSPN (Table S9). The SCFAs, derived from gut bacterial fermentation, have been considered as the most important bacterial metabolites affecting gut permeability and regulate intestinal and systemic inflammation in humans (Koh et al., 2016). We used the abundances of genes that encode key enzymes to indicate changes in the SCFAs genetically producing capacity (including acetic acid, propionate, and butyric acid) production pathways (Claesson et al., 2012). Most of the 54 HQMAGs carried *fhs* gene involved in the acetic acid biosynthesis pathway, regardless of the Guilds they belonged to (Table S9). For genes encoding the key enzyme propionyl-CoA:succinate-CoA transferase and propionate CoA transferase (*pst* and *pct*) in the propionate synthetic pathway, which has been reported to be negatively associated with glycemic control (Sanna et al., 2019; Tirosh et al., 2019), significantly much more HQMAGs in Guild 2 than that in Guild 1 harbored *pst* or *pct* (*pst*: 14.29 in Guild 1 vs 78.79% in Guild2, Fisher’s exact test *P* = 4.01×10 ^-6^ ; *pct*: 23.81% in Guild 1 vs 78.79% in Guild 2, Fisher’s exact test *P* = 1.59×10 ^-4^) (Figure 5D and Table S9). Conversely, only the HQMAGs in Guild 1 had *but* and *buk* genes, which are the predominate terminal genes for the butyrate biosynthetic pathways in gut microbiota (Figure 5D and Table S9). Moreover, we also assessed the change in endotoxin (LPS) biosynthesis, which is the major antigen from the gut microbiome. Lipid A, the amphipathic LPS glycolipid moiety, stimulates the immune system by tightly binding to Toll-like receptor 4 (Zhang et al., 2021). We showed that most of the HQMAGs in Guild 2 (78.79%) carried the genes related to Lipid A biosynthesis, but only two HQMAG in Guild 1 had these genes (Figure 5D and Table S9). In terms of antibiotic resistance genes (ARGs), 8 HQMAG s in Guild 1 encoded 14 ARGs and 29 genomes in Guild 2 encoded 112 ARGs (Figure 5D and Table S9). Taken together, FMT induced increase of Guild 1 that had the beneficial capacity for butyrate production and decrease of Guild 2 that could produce more antigen load, which contribute to the reduced low-grade, systemic and chronic inflammation.

### Matched enterotype between FMT transplants and recipients linked to better improvement of DSPN

In the FMT studies, it is always an interesting question whether the resemblance between transplants of gut microbiota from the healthy donors and/or the original gut microbiota in the recipients could be linked to the therapeutic efficacy. Here, we clustered all the samples, including 27 transplants and 150 samples from patients during FMT, into two enterotypes (C1 and C2) based on Jaccard distance of their microbiota at HQMAGs-level (Figure 6A). There were 19 transplants and 17 original gut microbiota in the recipients (samples of the patients at 0D) classed to C1, and 8 transplants and 15 original gut microbiota in the recipients classed to C2 (Figure 6B). Firstly, in order to test whether the specific transplant enterotype can affect therapeutic efficacy of FMT, we subjected the patients in two subgroups based on enterotypes of the transplants they received. TCSS scores in these two subgroups both showed a significant reduction at 84D after FMT, but there was no significant difference between the two subgroups (Figure S8). Next, we resubjected the patients into transplant-recipient matched or unmatched groups based on whether their original gut microbiota and transplants belonged to the same enterotype. Notably, the abundance of Guild 1 that negatively correlated with TCSS score, was significantly higher in the transplant-recipient matched than that in unmatched group from 56D to 84D after FMT (Figure 6C). Meanwhile, the abundance of Guild 2 that positively correlated with TCSS score, only significantly decreased in transplant-recipient matched group at 3D and kept on lower level from 28D to 84D after FMT, which was significantly lower than that in unmatched group (Figure 6C). Moreover, transplant-recipient matched group had significantly lower TCSS score than unmatched group at 84D (Figure 6D). These results suggest that matched enterotype between transplants and recipients in FMT may introduce better alteration in gut microbiota of the patients and induce better improvement of the symptoms and signs of DSPN.

**Figure 6.**
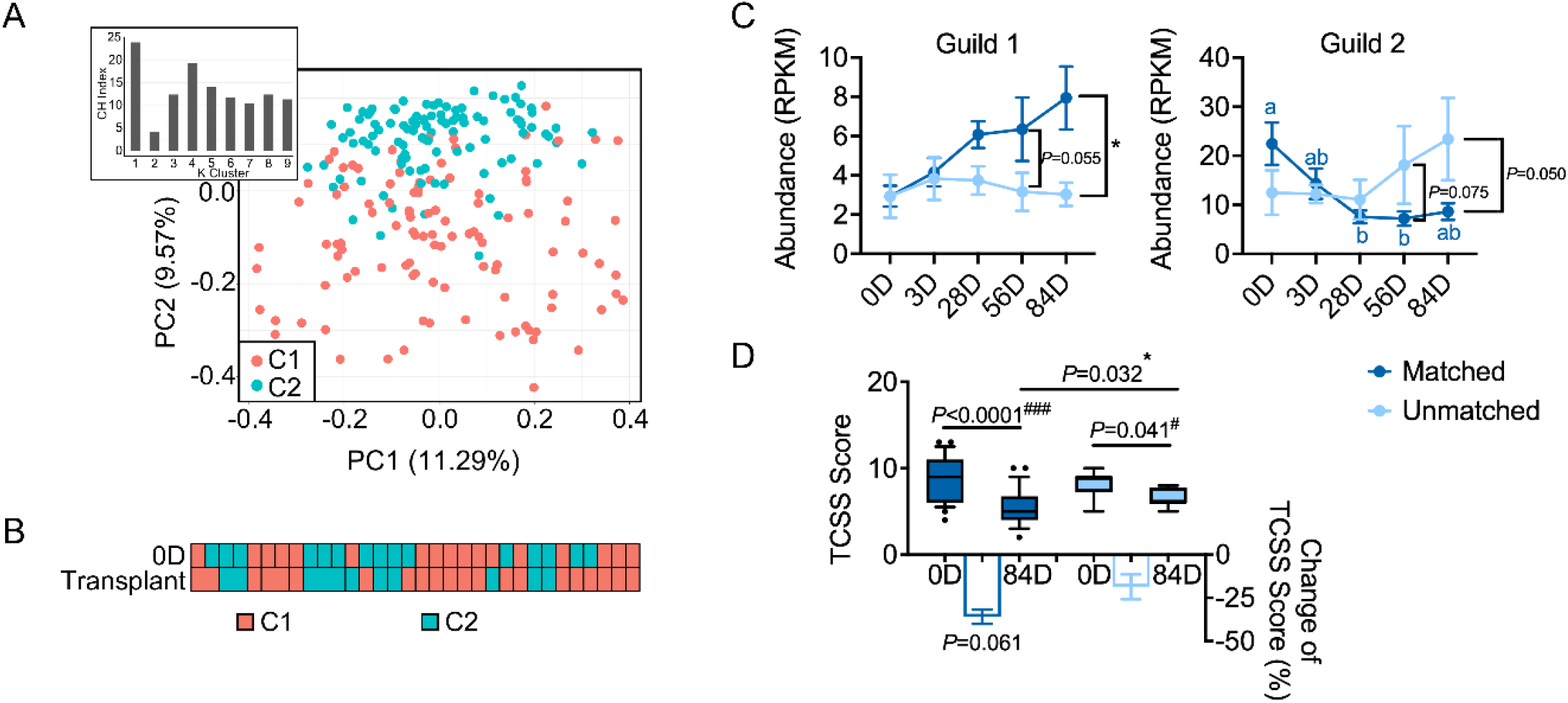
Matched enterotype between FMT transplants and recipients linked to better improvement of DSPN. **(A)** Classification of two enterotypes among all samples based on Jaccard distance and complete linkage. The samples included all the transplants, FMT group in RCT study and Placebo group in the post-RCT study of transplantation with fecal microbiota of healthy donors. *Calinski-Harabasz* (CH) index was used to assess the optimal number of clusters. (**B**) The enterotype of baseline gut microbiota of recipients paired with their transplants. (**C**) The abundance of Guild 1 and Guild 2 in matched and unmatched groups. The samples from FMT group in RCT study and samples from Placebo group in the following trial of transplantation with transplants from healthy donors were analyzed together. Matched group indicates the baseline gut microbiota of recipients and their transplants belong to the same enterotype (n = 24). unmatched group indicates the baseline gut microbiota of recipients and their transplants belong to the different enterotypes (n = 8). 0D indicates before transplantation, 3D, 28D, 56D and 84D indicate 3 days, 28 days, 56 days and 84 days after transplantation with transplants from healthy donors, respectively. Data are presented as the mean ± s.e.m. Mann-Whitney U test was used to analyze differences of abundance of Guild 1 or 2 between matched and unmatched groups, \**P* < 0.05. Friedman test followed by Nemenyi post-hoc test was used to compare the different timepoints within the same group, and values of each group with same letters are not significantly different (*P* > 0.05). (**D**) Changes of TCSS score in matched and unmatched groups. In the box plots, the line in the middle of the box is plotted at the median, and the inferior and superior limits of the box correspond to the 25^th^ and 75^th^ percentiles, respectively. The whiskers correspond to the 10^th^ and 90^th^ percentiles, and outliers are denoted. Mann-Whitney U test was used to analyze differences between matched and unmatched groups. * *P* < 0.05. Wilcoxon matched-pairs signed rank test was used to analyze each pairwise comparison 0D and 84D within each group. ^#^ *P* < 0.05 and ^##^ *P* < 0.01. The bars represent the mean change from the baseline value per group, with the corresponding s.e.m. Mann-Whitney U test was used to analyze differences in the changes between the matched and unmatched groups (intergroup changes).

## DISCUSSION

Our study shows that dysbiosis of the gut microbiota in patients contributed to the development of peripheral neuropathy, and modulation of the composition and function of gut microbiota via FMT alleviated neuropathic symptoms and improved sensory and motor nerve function in patients with DSPN.

We found the evidence of a causal relationship between the overall gut microbiome and peripheral nervous system disorders via conducting FMT from human to animal and from human to human. Although gut microbiota has been reported to have a role in the pathophysiology of neurological disorders related to enteric nervous system and central nervous system (Cryan et al., 2020; Joly et al., 2021; Morais et al., 2020), the knowledge of the relationship between the gut microbiome and the peripheral nervous disorders is still scanty. Here, we found that compared with control cohorts, patients with DSPN had a distinct gut microbiota composition. Transplanting the gut microbiota from patients with DSPN into the genetically diabetic mice, which received an antibiotic cocktail treatment in advance, induced gut barrier dysfunction, higher antigen load and severer systemic inflammation and aggravated peripheral neuropathy. Moreover, our RCT FMT trial showed that the transplanted gut microbiota from healthy donors was the only trigger for improving the nerve function and neuropathic symptoms in patients with DSPN, indicating a causal role of the gut microbiota(Cartwright, 2011). More importantly, the changes of gut microbiome in patients with DSPN induced by FMT occurred before the alleviation of the symptoms, supporting that the changes in the gut microbiota induced by FMT contributed to the improvement of DSPN rather than a mere consequence after severity had been alleviated.

To search for the key bacteria that may mediate the protective effect of the whole gut microbiota in DSPN, we adopted the genome-centric and guild-based approach(Wu et al., 2022; Wu et al., 2021) for analyzing the microbiome data before and after FMT. We did de novo assembly of high-quality genomes from the metagenomic datasets. Using these metagenome-assembled genomes as basic variables allows us to analyze the microbiome data at a resolution much higher than species or any higher taxa(Zhang and Zhao, 2016). We established an ecological network with genomes which were significantly correlated with the primary outcome TCSS score. This allows us to explore how health relevant bacteria in the gut ecosystem interact with each other and form higher level organizations to exert functions(Wu et al., 2021). These genomes turned out to be organized into two competing guilds, one beneficial and one detrimental. The beneficial Guild 1 had much higher genetic capacity in butyrate production and much less genes in synthetic pathway of endotoxin than the detrimental Guild 2. Such a seesaw-like network structure with two competing guilds has been reported as a core microbiome signature associated with various chronic diseases(Wu et al., 2022). Here, FMT effectively increased beneficial Guild 1 and decreased detrimental Guild 2 in the patients with DSPN, leading to a similar abundance of the two guilds to the transplants from healthy donors. Such changes were consistent with the alleviation of DSPN. More importantly, when the transplant and recipient microbiota were of the same enterotype, FMT had better efficacy than the unmatched. Patients receiving transplants with matched enterotype had more increase of Guild 1 and more decrease of Guild 2. This points not only to the mediating role of the two competing guilds in successful FMT but also the importance of the resemblance of the gut microbiota between donor and recipient for improving the therapeutic efficacy of FMT(Holvoet et al., 2021; Kim et al., 2020; Olesen and Gerardin, 2021).

Our study provides new insights on the potential mechanisms of the effectiveness of FMT in alleviating DSPN. Nerve dysfunction and neuronal cell death in DSPN results from a complex myriad of events that are triggered by the metabolic imbalances associated with diabetes (Feldman et al., 2017), and improvement of glycaemic control and life style are used as conventional treatment to alleviate the symptoms in patients with DSPN. However, glucose and lipid metabolism remained similar between the FMT and control groups of patients throughout our study, suggesting that the alleviations of nerve function and neuropathic symptoms in patients with DSPN after FMT may not be mediated via directly improving glycaemic response. The increased capacity for butyrate production and decreased for endotoxin from the two competing guilds of the gut microbiota may explain the beneficial effect of the gut microbiome after FMT on DSPN. Because reduced production of endotoxin from gut microbiome may reduce antigen load and decrease inflammation (Pan et al., 2018). Butyrate produced by gut microbiota is critical for maintenance of epithelial lining and gut barrier integrity via multiple intracellular processes, including providing essential energy source for colonocytes, activating of the transcription factor hypoxia-inducible factor-1 (HIF-1) to increase the expression of components of intestinal epithelial cell tight junctions, stimulating of mucus production etc. (Xiao et al., 2020). Improved gut barrier function may lead to further reduction of transfer of endotoxin from the gut to the circulation (Pan et al., 2018). More importantly, butyrate is reported to be positively correlated with the pain improvement following FMT and modulate gene expression and immune cells in the peripheral never system (PNS) (Bonomo et al., 2020). Though we did not investigate the inflammation in PNS, we found a significant alleviation of systemic inflammation as evidenced by significant decrease of LBP, TFN-α and IL-6 in the patients with DSPN after FMT. In the pathologic process of DSPN, hyperglycaemia, dyslipidaemia, and/or insulin resistance promote activation of the polyol, advanced glycation end products, protein kinase C, poly (ADP-ribose) polymerase, and hexosamine pathways as well as well loss of insulin signaling, which culminates in deleterious effects on mitochondrial function and gene expression along with oxidative stress and inflammation (Dewanjee et al., 2018; Sloan et al., 2021). Among all these complicated pathological changes in both type1 and type 2 diabetes, systemic inflammation may play a critical role in the development of DSPN. Proinflammatory cytokines not only enhance existing inflammatory and immune responses but also increase cellular oxidative/nitrosative stress, promoting even more neuronal damage in experimental models of DSPN (Schlesinger et al., 2019; Vincent et al., 2013). Systemic inflammation is involved in many complexed metabolic and immune processes and conditions, it is not specific to DSPN. The molecular mechanism of the gut microbiota in the development of DSPN need further investigation.

Taking the gut microbiota further into the investigation of DSPN may help us reveal novel mechanisms in its pathogenesis and may provide a new target for developing effective clinical therapies for this common, and pernicious unmet medical need in patients with diabetes mellitus.

## Data Availability

The data that support the findings of this study are available from the corresponding author upon request. Metagenomic sequence data have been submitted in DDBJ (http://www.ddbj.nig.ac.jp) with accessions number DRA009982. Amplicon sequence data have been submitted in NCBI SRA (https://www.ncbi.nlm.nih.gov/sra) with accessions number SRP379656, SRP278004 and SRP272175.

https://www.ncbi.nlm.nih.gov/sra

## ACKNOWLEDGEMENTS

We thank Miao Wang from China Microbiota Transplantation System for offering the WMT and placebo data sources. We acknowledge a computing facility award for use of the Pi cluster at Shanghai Jiao Tong University. This work was supported by the following funding supports: National Natural Science Foundation of China (81970705, 81871091, and 31922003).

## AUTHOR CONTRIBUTIONS

H. Y., C. Z., F. Z. and L. Z. designed this study. J. Y., X. S., J. S., Y. F., X. D., and S. T. recruited and supervised the participants and performed all clinical procedures. F. H., J. Y., and X. S. completed the animal experiment. F. Z., P. L. and B. C. provided WMT and the related protocol. W. W., H. Z., L. C., X. W. and Y. L. completed the delivery of FMT. X. Y., Y. Z., X. W., and Y. L. performed preparation, processing and sequencing of fecal samples. R. Z. analyzed the 16S rRNA sequencing data. G. W., and C. Z. analyzed the metagenomic sequencing data. J. Y., X. Y., X. S., X. W., L. W. and Q.Y. supervised operation of ELISA. C. Z., J. Y. and G. W. performed statistical analysis and generated the figures and tables. C. Z., H. Y., L. Z., J. Y., X. Y. and G. W. prepared the manuscript. All authors approved the manuscript.

## STAR ★ METHODS

Detailed methods are provided in the online version of this paper and include the following:

## KEY RESOURCES TABLE

## CONTACT FOR REAGENT AND RESOURCE SHARING

## EXPERIMENTAL MODEL AND SUBJECT DETAILS

- Clinical Study
- Animal study with db/db Mice

## METHOD DETAILS

- Questionnaire
- Neurophysiological Examination
- Immunofluorescence and Immunohistochemistry
- Serum ELISA
- Gut Microbiome Analysis

## QUANTIFICATION AND STATISTICAL ANALYSIS

- Statistical Analysis of clinical data
- Statistical analysis of gut microbiota data

## DATA AND SOFTWARE AVAILABILITY

## KEY RESOURCES TABLE

| REAGENT or RESOURCE | SOURCE | IDENTIFIER |
| --- | --- | --- |
| Antibodies |  |  |
| Human ZO-1 Rabbit Polyclonal antibody | Proteintech | 21773-1-AP |
| Human Anti-Claudin-1 antibody | Abcam | Ab211737 |
| Human Anti-Claudin 4 antibody | Abcam | Ab53156 |
| Goat anti-rabbit cyanine 3 (Cy3) (red) | Boster Biological Technology | BA1032 |
| Mouse Anti-ZO-1 antibody | Servicebio | GB111981 |
| Mouse Anti-Claudin-1 antibody | Servicebio | GB11032 |
| Mouse Anti-Claudin 4 antibody | Proteintech | 16195-1-AP |
| Mouse Anti-NF 200 antibody | Proteintech | 18934-1-AP |
| Mouse Anti-MBP antibody | Abcam | Ab218011 |
| Mouse Anti-BDNF antibody | Abcam | Ab108319 |
| Goat Anti-Rabbit IgG | Servicebio | GB23303 |
| Critical Commercial Assays |  |  |
| Human TNF-α Immunoassay ELISA kit | R&D Systems Minneapolis | HSTA00E |
| Human IL-6 ELISA Kit | Shanghai Jianglai Industrial Limited by Share Ltd. | JL14113 |
| Human LBP ELISA kit | Hycult®Biotec | HK315 |
| Mouse LBP ELISA Kit | Elabscience | E-EL-M2686c |
| Mouse IL-6 ELISA Kit | Elabscience | E-EL-M0044c |
| Mouse TNF-α ELISA Kit | Elabscience | E-EL-M3063 |
| PowerFecal® DNA Kit | QIAamp | 12830-50 |
| Deposited Data |  |  |
| NCBI | https://www.ncbi.nlm.nih.gov/sra | SRP379656,<br>SRP278004,<br>SRP272175 |
| ENA | https://www.ebi.ac.uk/ena/browser/home | PRJEB53551 |
| Software and Algorithms |  |  |
| R project (version 3.5.3) | CRAN | https://cran.r-project.org/ |
| blockrand (version 1.5) in R project | https://cran.r-project.org/mirrors.html |  |
| MASS (version 7.3-51.4) in R project | https://cran.r-project.org/mirrors.html |  |
| ade4 (version 1.7-15) in R project | https://cran.r-project.org/mirrors.html |  |
| QIIME 2 version 2019.10 | https://qiime2.org |  |
| CoverM v0.6.1 | https://github.com/wwood/CoverM |  |
| Resistant gene identifier 5.2.1 | https://card.mcmaster.ca/analyze/r<br>gi |  |

## CONTACT FOR REAGENT AND RESOURCE SHARING

Further information and requests for resources and reagents should be directed to and will be fulfilled by the Lead Contact, Huijuan Yuan

## EXPERIMENTAL MODEL AND SUBJECT DETAILS

### Clinical Study

The studies were approved by the Medical Ethics Committee of Henan Provincial People’s Hospital and written informed consent was obtained from all participants.

### Comparation of gut microbiota among subjects with normal glucose level and DM patients with or without DSPN

Firstly, we recruited 27 patients with DSPN. Patients were recruited if they were (1) type 1 or type 2 diabetes with DSPN. The diagnosis of DSPN was according to the criteria recommended by 2017 American Diabetes Association(Pop-Busui et al., 2017). According to the criteria, the patients had (i) at least one abnormal symptoms (including local or distal numbness, pain, formication, and tingling) and signs (including pinprick sensation, temperature sensation, vibration perception, proprioception, 10-g monofilament, and ankle reflexes), with TCSS score > 5 or VAS score⩾ 4; (ii) excluded other disease caused peripheral neuropathy; (2) aged 18-70 years old; (3) with glycated haemoglobin (HbA1c) <11%; (4) responded poorly to conventional treatments for at least 3 months (84days). Besides, the conventional treatments include lifestyle modification, glucose control, and drug intervention; the patients responded poorly to conventional treatments means their TCSS scores decreased by < 3 and/or VAS scores decreased by < 25% as compared to that before treatment.

Patients were excluded if they (1) had a continuous antibiotic use history for > 3 days within 3 months prior to enrolment; (2) had any clinically significant or unstable mental or psychiatric illnesses or epilepsy; (3) had other causes of neuropathy, such as osteoarthritis, cervical lumbar diseases, connective tissue disease, peripheral vascular disease, tumor peripheral neuropathy, herpes zoster infection, abnormal thyroid function, or severe malnutrition; (4) had undergone gastrectomy, fundoplication, colostomy or other digestive system surgery; (5) had persistent vomiting or a suspected gastrointestinal obstruction; (6) had taken drugs that can cause peripheral neuropathy, such as isoniazid or furazolidone; (7) had severe cardiovascular and cerebrovascular diseases or liver, kidney and haematopoietic system diseases; (8) had alcoholism (drinking more than 5 times in one week, more than 100 g of spirits, 250 g of rice wine or 5 bottles of beer); (9) were pregnant; (10) had a physical disability or self-care disability or were unable to recall clearly and answer questions due to any other reasons; or (11) lacked the time to take part in this project. Written informed consent was obtained from all participants. The feces were collected and stored at -80°C.

From our previous clinical trial with the DM patients published by Xinru Deng et al (Deng et al., 2022), we selected 30 age-and sex-matched patients without DSPN (had no DSPN symptoms and TCSS score⩽5, n=30) and used their baseline data (before drug treatment). The data of the subjects with normal glucose level (n=29) was obtained from our previous cohort study published article by Yuanyuan Fang et al (Fang et al., 2021).

### Randomized controlled trial

This randomized, double-blind, placebo-controlled pilot clinical trial was registered in the Chinese Clinical Trial Registry (ChiCTR1800017257), and performed from August 2018 to February 2022. Written informed consent was obtained from all participants.

#### Sample Size Estimation

Because no clinical trials of FMT treating DSPN performed previously, the proof of concept study enrolled 39 subjects. No sample size calculation was performed.

According to this observed magnitude of changes (3.09 in FMT v.s. 0.6 in Placebo) and standard deviation (2.4 in FMT and Placebo) of TCSS scores from our RCT study, the power calculation estimated that 36 participants (FMT: Placebo=2:1) would provide α=0.05, 1-β=0.8, and there would be approximately 40 subjects participate coupled with a dropout rate of 10%. Then the sample size of our RCT study was similar with this calculated value.

#### Study Subjects

We recruited DSPN patients with the same inclusion and exclusion criteria as the above description. And the participant flow was shown in Figure S3.

#### Clinical trial of FMT

During the 14-day run-in period, all antidiabetic medications except insulin were withdrawn, all conventional treatments for DSPN was stopped and other non-diabetic drugs which long term use before FMT remained unchanged to avoid potential confounding effects on the gut microbiota. At the same time, all patients were given dietary guidance according to the Diabetic Diet Guidelines in Chinese (2017). At baseline, all participants’ characteristics were assessed comprehensively, peripheral blood was collected for chemical and biological analyzes, and stool samples were collected for gut microbiota analysis. Upon successful completion of the 14-day run-in, participants were randomly assigned to the intervention or control group in a 2:1 ratio and followed up for 84 days.

Randomization codes were determined by a computer-generated random sequence (random sequence generation by blockrand (version1.5) package was performed using R project). In order to double-blind, the randomization and the placebo was conducted by the operator of nonprofit China fmtBank who did not participate in the enrolment and transplantation process. All the doctors and operators were blinded, and unblind was given by the nonprofit China fmtBank after the patients completed the last visit (84 days).

The primary indicator of the study was the change of TCSS score at 84D compared with baseline and the secondary indicators included the change of VAS score, HAMA score, HAMD score, PSQI and WHOQOL-BREF score, NCV and CPT at 84D compared with baseline. All adverse events were recorded and described according to the Common Terminology Criteria for Adverse Events (CTCAE). The mechanism indexes were the structure and function of gut microbiota, inflammatory cytokine and gut barrier integrity.

#### Donor selection and transplants preparation

Donors were selected and screened by the nonprofit China fmtBank. The criteria for FMT donor screening included eight aspects: age, physiology, pathology, psychology, veracity, time, living environment and recipients. The ages of the donors ranged from 18-24 years, and all of them were evaluated for physiological status, such as for body growth, body mass index, sleep quality, daily habits, diet, physical exercise and regular bowel habits. The detailed protocol was reported in our previous papers (Ding et al., 2019; Zhang et al., 2020).

The methodology of FMT used in the present study was recently designated washed microbiota transplantation (Zhang et al., 2020), which was performed using automated instruments (GenFMTer, FMT Medical, Nanjing, China) in a biosafety level 3 laboratory at Nanjing Medical University. Information regarding the donors and laboratory processes was recorded by the China Microbiota Transplantation System (CMTS). We used the one-hour protocol, which requires all steps from defecation to storage of the microbiota suspension in a -80°C freezer to be conducted within one hour. The enriched microbiota was collected according to the washed preparation protocol reported by recent report (Zhang et al., 2020) and the panel consensus report (2020). The placebo, which was identical in appearance, form, color and size to the frozen microbiota, was composed of saline with food grade coloring pumpkin powder and purple potato powder (total 13.2g for the total administration per day).

#### FMT delivery and follow-up

The methods used for delivering the washed microbiota were described in our previous report (Dai et al., 2019). Briefly, patients were prepared according to the method used for routine gastroscopy and colonoscopy. Patients underwent mid-gut transendoscopic enteral tubing (TET) for insertion of a 2.7 mm outer-diameter tube under anaesthesia (Long et al., 2018). All patients underwent colonoscopy to exclude complicated intestinal diseases, and six mucosal biopsies were obtained at the junction of the descending and sigmoid colons in both FMT (n=6) and placebo (n=5) group. Tissues were stored in liquid nitrogen for further immunofluorescence testing. The frozen fecal microbiota or placebo from China fmtBank was thawed to 37°C in a water bath. Total 150 mL rewarming suspension with 5 × 10^13^ of bacteria as the total dose per day was delivered into intestine via mid-gut tube. Total two doses were delivered within two days. The patients were blinded to the infusion provided by operators. Patients maintained a seated position for 30 min and fasting state for 2 h after transplant. The microbiota from the same batch was infused through the mid-gut TET tube on the second day.

All patients were evaluated at scheduled follow-up visits at 3 days (3D), 28 days (28D), 56 days (56D) and 84 days (84D) after FMT. The antibiotics, probiotics, and yogurt were unable to be taken throughout the follow-up. At each visit, patients underwent examination of the TCSS, VAS, HAMA, HAMD, PSQI and WHOQOL standards, and dietary guidance was given at the same time, all of them were conducted by one experienced and constant investigator. Peripheral blood was collected and serum samples were stored at -80°C. The stool samples were collected and stored at -80°C. Moreover, the levels of NCV and CPT were also assessed at baseline (0D) and at the end point (84D). Most notably, for the last visit (84D), all the above operating procedures were accomplished before unblinding. All adverse events described according to the Common Terminology CTCAE were submitted to the CMTS for long-term monitoring. The grade refers to the adverse event severity. The CTCAE displays grades 1–5 with clinical descriptions of adverse event severity based on the guidelines (Table S4).

### Post-RCT study

After finishing the RCT study, all patients in the placebo group of the above RCT received transplants made from fecal microbiota of the healthy donors, following with the same protocol of the RCT study for treatment, follow-up, test of clinical parameters and samples collection.

### Animal study with db/db Mice

#### Preparation of transplants from DSPN patients and NG subjects

In the anaerobic operating chamber (80% N2:10% CO2:10% H2,Don Whitley Scientific, UK), an equal amount of frozen feces from DSPN patients (n=5) or NG subjects (n=5) were melted and mixed at 37° C. Each mixed fecal material (1 g) was diluted in 50 mL of a sterile Ringer working buffer (9 g/L of sodium chloride, 0.4 g/L of potassium chloride, 0.25 g/L of calcium chloride dehydrate and 0.05% (w/v) L-cysteine hydrochloride). The diluted fecal materials were suspended by vortex for 5 min and then settled by gravity for 5 min. The clarified supernatant was transferred to a clean tube, and an equal amount of 20%(W/V) skim milk was added. The transplant was prepared fresh on the day of the transplantation experiment, and the rest was stored at -80°C until the inoculation.

#### Animal study

All animal experimental procedures were approved by the Committee of Animal Experimental Center of Zhengzhou University (ZZU-LAC20211015[10]), and were conducted according to the committee’s guidelines. Teen-week-old SPF male BKS-DB (Lepr)(db/db) mice were purchased from Jiangsu Jijiyukang Biotechnology Co., LTD. (Certificate No. SCXK(Su)2018-0008) and kept under SPF environment at the Zhengzhou University Experimental Animal Center. The mice were fed with a sterilized normal chow diet (3.9kcal/g; HUANYU BIO, GB 14924.3-2010) and housed in a room maintained at 22 ± 2 ° C with humidity of 50%±10%, with a 12 h light/dark phase cycle (lights on at 07:00 am).

After one-week of acclimation, all the db/db mice were treated with vancomycin (0.5 g/L), neomycin sulfate (1g/L), ampicillin (1g/L) and metronidazole (1g/L) in drinking water. After 2 weeks with the antibiotic cocktail treatment, the mice were then randomly assigned to 2 groups: (i) M-DSPN group (n = 6), the db/db mice inoculated with transplant from DSPN patients; (ii) M-NG group (n = 6), the db/db mice inoculated with transplant from NG subjects. The oral gavage was performed once a day for the first 3 days, and strengthened once every 3 days. After 3 weeks, the feces were collected and mechanical sensitivity, thermal sensitivity and motor nerve conduction velocity were used to measure neuropathic indicators. Then the mice were scarified, and the blood samples and tissues of clone, dorsal root ganglion and sciatic nerve were collected. All the samples were stored at -80°C.

## METHOD DETAILS

### Questionnaire

#### Toronto Clinical Scoring System (TCSS)

The severity of DSPN was assessed with TCSS (Arumugam et al., 2016), which including the scores of symptoms, reflexes and sensory tests. The symptom scores contain pain, numbness, tingling and weakness of the lower limb; ataxia and symptoms of upper limb. The deep tendon reflexes contain knee and ankle reflexes. The sensory tests contain pinprick sensation, temperature sensation, light touch, vibration and position, which were performed on the first toe. The TCSS was investigated by one experienced and constant investigator.

Symptom scores: score of 0 represents absent, score of 1 represents present. Reflex scores: score of 2 represents absent, score of 1 represents reduced, score of 0 represents normal. Sensory test scores: score of 1 represents abnormal, score of 0 represents normal. The higher the overall score, the more severe the symptoms.

#### Visual Analogue Scale (VAS)

The severity of neuropathic pain of the patients were evaluated by VAS (Petersen et al., 2021). 10 cm of Horizontal line was drawn on the paper, one end of the horizontal line is 0, indicating no pain; the other end is 10, indicating severe pain; the middle part shows different levels of pain. The patients were asked to mark the level of any form of pain on the line according to their feelings. A score of 1–3 indicated mild pain and 4–10 indicated moderate/severe pain.

#### Hamilton Anxiety Scale (HAMA)

Anxiety was evaluated by means of the Hamilton Anxiety Scale (HAMA (Zhao et al., 2021)). HAMA includes 14 items; each item is scored from 0 to 4, with a higher score reflecting more severe anxiety.

#### Hamilton Depression Rating Scale (HAMD)

Depression was assessed by means of the Hamilton Depression Scale (HAMD (Zhao et al., 2021)). It consists of 17 items; each item is scored from 0 (not present) to 7 (severe), with a higher total score indicating more severe depression.

#### Pittsburgh Sleep Quality Index (PSQI)

The sleep quality was evaluated by PSQI (Buysse et al., 1989), which mainly consists of 19 self-rated questions, which assess a wide variety of factors relating to sleep quality, including estimates of sleep duration and latency and of the frequency and severity of specific sleep-related problems. These I9 items are grouped into seven component scores, each weighted equally on a 0-3 scale. The seven component scores are then summed to yield a global PSQI score, which has a range of 0-21; higher scores indicate worse sleep quality.

#### Brief table of the World Health Organization’s Quality of Life (WHOQOL-BREF)

The quality of life was assessed by WHOQOL-Bref (Chiu et al., 2006; Saxena et al., 2001), which containing 26 items and are represented by the most suitable one item for each of 24 WHOQOL-100 facets. The 24 facets or items are further categorized into four domains: physical capacity (7 items), psychological well-being (6 items), social relationships (3 items), and environment (8 items). Each item uses a scale from 1 to 5, with a higher score indicating a higher quality of life. Domain scores are calculated by multiplying the mean of all facet scores included in each domain by a factor of 4, and potential scores for each domain vary from 4 to 20 (e.g., score of social relationships = ((Q20 + Q21 + Q22)/3) * 4).

#### Neurophysiological Examination

##### CPT

The Neurometer^R^CPT detector (Neurotron Inc., Baltimore, USA) was used to test CPT, which was operated by one experienced and constant investigator following the standard protocol of the equipment. The patient took a seat position, fully exposed the inspection site and the room temperature kept at 24±1 degrees. There were four test sites, the distal phalanx of index finger on the both dorsal hands and the distal phalanx of hallux on the both dorsal feet. The test points were stimulated with 2000Hz, 250Hz and 5Hz sine wave current. According to the manufacturer, each result below or above the reference range was defined as hyperesthesia or hypoesthesia, respectively.

##### Nerve conduction velocity

For clinical studies, electromyography (MEB-9400C, Nihon Kohden Corporation, Tokyo, Japan) was used to assess the MNCV and SNCV, which was operated by one experienced and constant investigator following the standard protocol of the equipment. The patient took a supine position, fully exposing the test site and surface electrodes were used to stimulate the nerve. The patient’s limb temperature was maintained at 32 degrees and the room temperature was kept at 28-30 degrees.

For mice study, nerve conduction velocity (NCV) was measured as described previously (Goss et al., 2002) with slight modifications. Mice were anesthetized with 10ml/kg chloral hydrate and electrodes were placed at the acrotarsium and sciatic notch. Nicolet EDX electromyography instrument was used to record simultaneous electromyography during electrical stimulation, and the motor nerve conduction velocity (MNCV) was calculated.

##### Mechanical and Thermal Sensitivities

For mice study, mechanical allodynia was assessed using calibrated Von Frey filaments (Stoelting) and thermal hyperalgesia was assessed by a thermal stimulation meter (YLS-6B) according to published methods (Fan et al., 2020).

### Immunofluorescence and Immunohistochemistry

#### Immunofluorescence analysis of the tight junction proteins in colon of patients

Frozen sections (4 μm thickness) were prepared from colon tissue from patients with a freezing microtome (Cryotome E, Thermo, MA, USA). Slides were blocked with diluted goat serum and then incubated with primary antibodies against ZO-1 (1:100), Claudin-1 (1:100), and Claudin-4 (1:100) at 4°C overnight. After being washed with PBST, the sections were incubated with goat anti-rabbit cyanine 3 (Cy3) (red) secondary antibodies (Boster Biological Technology, CA, USA). Nuclei were visualized with 4-6-diamidino-2-phenylindole-2 HCl (DAPI, Beyotime, Jiangsu, China). The stained slides were then scanned by scanning confocal microscopy (BX53, Olympus, Tokyo, Japan).

For each immunofluorescence marker, we assessed a single 5-mm section. Three regions of interest (ROIs) were manually selected from each 5-mm section per marker per sample. Each ROI measured 0.5 × 0.5 mm ^2^, and the density of each marker plotted for each time point represents the mean of the four ROIs per mm^2^. The immune infiltrate density was quantified digitally by using Image-Pro Plus 6.2 (Media Cybernetics Inc., Rockville, MD, USA).

#### Immunohistochemistry analysis in mice study

##### Intraepidermal nerve fiber density measurement

The plantar skin of the mice was drilled horizontally with a skin biopsy instrument with a diameter of 2mm. The plantar skin including epidermis and dermis was cut out by ophthalmic scissors and directly fixed with 4% paraformaldehyde for subsequent immunohistochemistry. Intraepidermal nerve fibre density (IENFD) was measured using PGP9.5 antibody staining in a blinded fashion. Sections were restrained with eosin (Sigma-Aldrich Eosin Y solution HT110316) to depict the fiber crossing at the dermoepidermal junction. IENFD was calculated (as fibres/mm) by the number of complete baseline crossings of nerve fibres at the dermo-epidermal junction (Chandrasekaran et al., 2019).

##### Dorsal root ganglion and sciatic nerve

The paraffin-embedded sections of dorsal root ganglion, sciatic nerve tissue and colon tissue samples were deparaffinized by xylene and dehydrated by different concentration of alcohol solutions and then were treated with citrate or EDTA buffer for antigen retrieval. 3 % H2O2 was used to block endogenous peroxidase for 15 min, followed by diluted goat serum to reduce nonspecific staining for 30 min. For dorsal root ganglion and sciatic nerve tissue, the sections were incubated with anti-NF 200 (1:300), anti-MBP (1:1000) and anti-BDNF (1:500) primary antibodies at 4 °C overnight.

##### Tight junction proteins in colon

The colon tissue samples the sections were incubated with anti-claudin-1 (1:600), anti-claudin-4 (1:200) and anti-ZO-1 (1:300) primary antibodies at 4 °C overnight. After incubation and subsequent washing, the secondary antibodies corresponding to the primary antibody were added to the samples and incubated at room temperature for 50 min. After washing three times with PBS, the slices were stained with diaminobenzidine (DAB). Images were obtained using a microscope and positive DAB-stained areas were calculated by the ImageJ software.

### Serum ELISA

For clinical studies, commercially available ELISA kits were used to determine serum levels of LBP (HK315, Hycult®Biotech, UDEN, Netherlands), TNF-α (HSTA00E, QuantikineHS, R&D Systems Minneapolis, MN, USA) and IL-6 (JL14113, Shanghai Jianglai Industrial Limited by Share Ltd., shanghai, China). The minimum detectable concentrations for LBP, TNF-α and IL-6 were 4.4 ng/mL, 0.011 pg/mL and 3.12 pg/mL, respectively.

For mice study, commercially available ELISA kits were used to determine serum levels of LBP (E-EL-M2686c, Elabscience, Wuhan, China), TFN-α (E-EL-M3063, Elabscience, Wuhan,China) and IL-6 (E-EL-M0044c, Elabscience, Wuhan, China). The minimum detectable concentrations for LBP, TNF-α and IL-6 were 3.13 ng/mL, 7.81pg/mL and 31.25 pg/mL, respectively.

### Gut Microbiome Analysis

#### DNA extraction

Genomic DNA was extracted from human and mice feces using the QIAamp PowerFecal Pro DNA Kit (QIAGEN, USA, 51804).

#### 16S rRNA gene V3-V4 region sequencing

PCR targeting the V3-V4 region of the 16S rRNA gene with primers Forward [5’-CCTACGGGNGGCWGCAG -3’] and Reverse [5’-GACTACHVGGGTATCTAATCC -3’] (Klindworth et al., 2013). The subsequent amplicon sequencing was performed on a MiSeq platform to generate paired-end reads of 300 bp (Illumina, CA, USA).

The reads were analyzed using QIIME2 version 2019.7 (Bolyen et al., 2019). Then the adapters of the sequences were removed using the “cutadapt” plugin of QIIME2. DADA2 was used to obtain the abundance and representative sequences of amplicon sequence variants (ASVs) (Callahan et al., 2016). Afterwards, the ASVs of three batches were merged using the QIIME2. Representative sequences for ASVs were built into a phylogenetic tree using core-metrics-phylogenetic pipeline in QIIME2 and were assigned into taxonomy using Silva database (release 132) (Quast et al., 2013). All the samples were randomly subsampled to equal depths of 23154 reads prior to the following analysis.

α-diversity analysis and principal coordinate analysis (PCoA) were conducted using QIIME2 diversity plugins. Permutational multivariate analysis of variance test (PERMANOVA, 999 tests) was performed using R package vegan.

#### Shot-gun Metagenomic Sequencing

DNA was sequenced using an Illumina HiSeq 3000 at GENEWIZ Co. (Beijing, China). Cluster generation, template hybridization, isothermal amplification, linearization, and blocking of denaturation and hybridization of the sequencing primers were performed according to the workflow specified by the service provider (Liu et al., 2016). Libraries were constructed with an insert size of approximately 500 bp, followed by high-throughput sequencing to obtain 150 bp paired-end reads in the forward and reverse directions.

Trimmomatic (Bolger et al., 2014) was employed to 1) trim adapters 2) remove low-quality bases and 3) remove short reads less than 60 bp in length. Reads that could be aligned to the human genome (*Homo sapiens*, UCSC hg19) were removed (aligned with Bowtie2 (Langmead and Salzberg, 2012)). The numbers of high quality reads obtained for each sample are shown in Table S6.

*De novo* assembly was performed for each sample with IDBA_UD (Peng et al., 2012). The assembled contigs were further binned by using MetaBAT2 (Kang et al., 2019) with default parameters. CheckM (Parks et al., 2015) was used to assess the quality of the bins. Bins with completeness > 95%, contamination < 5% and strain heterogeneity < 0.05 were retained as high-quality draft genomes (Table S7). To improve the analysis, we also downloaded genomes from the HGG constructed by Samuel *et al* (Forster et al., 2019). The assembled high-quality draft genomes and the HGG genomes were dereplicated by using dRep (Olm et al., 2017) to obtain nonredundant genomes for further analysis (if the dRep cluster contained the draft genomes assembled from our dataset, we used the best genomes within the assembled genomes as the representative genomes of the clusters). The abundance of the genomes was calculated using CoverM v0.6.1 (https://github.com/wwood/CoverM) with parameters: --min-read-aligned-percent 90 --min-read-percent-identity 99. Taxonomic assignment of the genomes was performed by using GTDB-Tk (Chaumeil et al., 2019) (Table S8).

Prokka (Seemann, 2014) was used to annotate the nonredundant genomes. KofamKOALA (Aramaki et al., 2020) was used to assign KEGG orthologue IDs to the predicted protein sequences in each genome by HMMSEARCH against KOfam. Lipid A biosynthesis associated genes were identified based on the KEGG ontology (KO) ID. The overall structural changes in gut microbial function after FMT were represented by PCoA of Bray-Curtis distances using KEGG Ontology (KO), which revealed the significant changes in gut microbial function after FMT product introduction, corresponding to the strain-level analysis results. The protein sequences for formate-tetrahydrofolate ligase, propionyl-CoA: succinate-CoA transferase and propionate CoA transferase were obtained from the NCBI database with a text search. The protein sequences for 4Hbt, AtoA, AtoD, Buk and But were obtained from the IMG database as described previously (Vital et al., 2014). The predicted protein sequences in each genome were aligned to these sequences using BLASTP (best hit with E-value < 1e^-5^, identity > 80% and coverage > 70%). Antibiotic resistance genes were identified by Resistance Gene Identifier based CARD database (Alcock et al., 2020).

## QUANTIFICATION AND STATISTICAL ANALYSIS

### Statistical Analysis of Clinical Data

Statistical analyzes were performed using R project (version 3.5.3). Values are expressed as the mean ± s.e.m., median with interquartile range (IQR), or number. For clinical study, One-way ANOVA or Kolmogorov-Smirnov test was used to detect the differences in clinical parameters among the DSPN, DM and NG groups. Mann-Whitney U was used to detect the differences between DM and NG groups. During the RCT study, Mann-Whitney U Test, Student’s t-test (two-tailed) or Chi-squared test was used to detect the differences between FMT and placebo group; Paired t-test (two-tailed) or Wilcoxon matched-pair signed-rank test was used to analyze each pairwise comparison within each group. During the post-RCT study, One-Way RM ANOVA test was used to analyze differences between the three time point. For animal study, Student’s t-test (two-tailed) was used to compare the differences between the two groups. All the above statistical calculation methods were carried out by MASS (version 7.3-51.4) package or ade4 (version 1.7-15) package.

### Statistical Analysis of Gut Microbiota Data

Subject adjusted PCoA plot of the genomes based on Jaccard distance were performed with aPCoA package (Shi et al., 2020). Wilcoxon matched-pair signed-rank tests were used to analyze each pairwise comparison and performed using package MASS (version 7.3-51.4) in R project. MaAslin2(Mallick et al., 2021) was used to find genomes associated with TCSS score using linear mixed effect model with subject as random effect. The default significance cutoff adjusted *P* value < 0.25 was used in our study. Repeat measure correlation was used to calculated the co-abundance correlations between TCSS correlated genomes (Bland and Altman, 1995).

## DATA AND SOFTWARE AVAILABILITY

## REFERENCES

(2020). Nanjing consensus on methodology of washed microbiota transplantation. Chinese medical journal 133, 2330–2332.

Alcock, B.P., Raphenya, A.R., Lau, T.T.Y., Tsang, K.K., Bouchard, M., Edalatmand, A., Huynh, W., Nguyen, A.V., Cheng, A.A., Liu, S., et al. (2020). CARD 2020: antibiotic resistome surveillance with the comprehensive antibiotic resistance database. Nucleic acids research 48, D517–D525.

Aramaki, T., Blanc-Mathieu, R., Endo, H., Ohkubo, K., Kanehisa, M., Goto, S., and Ogata, H. (2020). KofamKOALA: KEGG Ortholog assignment based on profile HMM and adaptive score threshold. Bioinformatics 36, 2251–2252.

Arumugam, T., Razali, S.N., Vethakkan, S.R., Rozalli, F.I., and Shahrizaila, N. (2016). Relationship between ultrasonographic nerve morphology and severity of diabetic sensorimotor polyneuropathy. Eur J Neurol 23, 354–360.

Bland, J.M., and Altman, D.G. (1995). Calculating correlation coefficients with repeated observations: Part 1--Correlation within subjects. BMJ 310, 446.

Bolger, A.M., Lohse, M., and Usadel, B. (2014). Trimmomatic: a flexible trimmer for Illumina sequence data. Bioinformatics 30, 2114–2120.

Bolyen, E., Rideout, J.R., Dillon, M.R., Bokulich, N.A., Abnet, C.C., Al-Ghalith, G.A., Alexander, H., Alm, E.J., Arumugam, M., Asnicar, F., et al. (2019). Reproducible, interactive, scalable and extensible microbiome data science using QIIME 2. Nat Biotechnol 37, 852–857.

Bonhof, G.J., Herder, C., Strom, A., Papanas, N., Roden, M., and Ziegler, D. (2019). Emerging Biomarkers, Tools, and Treatments for Diabetic Polyneuropathy. Endocr Rev 40, 153–192.

Bonomo, R.R., Cook, T.M., Gavini, C.K., White, C.R., Jones, J.R., Bovo, E., Zima, A.V., Brown, I.A., Dugas, L.R., Zakharian, E., et al. (2020). Fecal transplantation and butyrate improve neuropathic pain, modify immune cell profile, and gene expression in the PNS of obese mice. Proc Natl Acad Sci U S A 117, 26482–26493.

Bulgart, H.R., Neczypor, E.W., Wold, L.E., and Mackos, A.R. (2020). Microbial involvement in Alzheimer disease development and progression. Mol Neurodegener 15, 42.

Buysse, D.J., Reynolds, C.F., 3rd, Monk, T.H., Berman, S.R., and Kupfer, D.J. (1989). The Pittsburgh Sleep Quality Index: a new instrument for psychiatric practice and research. Psychiatry Res 28, 193–213.

Callahan, B.J., McMurdie, P.J., Rosen, M.J., Han, A.W., Johnson, A.J., and Holmes, S.P. (2016). DADA2: High-resolution sample inference from Illumina amplicon data. Nat Methods 13, 581–583.

Cani, P.D., Depommier, C., Derrien, M., Everard, A., and de Vos, W.M. (2022). Akkermansia muciniphila: paradigm for next-generation beneficial microorganisms. Nat Rev Gastroenterol Hepatol.

Cartwright, N. (2011). A philosopher’s view of the long road from RCTs to effectiveness. Lancet 377, 1400–1401.

Chandrasekaran, K., Salimian, M., Konduru, S.R., Choi, J., Kumar, P., Long, A., Klimova, N., Ho, C.Y., Kristian, T., and Russell, J.W. (2019). Overexpression of Sirtuin 1 protein in neurons prevents and reverses experimental diabetic neuropathy. Brain 142, 3737–3752.

Chaumeil, P.A., Mussig, A.J., Hugenholtz, P., and Parks, D.H. (2019). GTDB-Tk: a toolkit to classify genomes with the Genome Taxonomy Database. Bioinformatics.

Chiu, W.T., Huang, S.J., Hwang, H.F., Tsauo, J.Y., Chen, C.F., Tsai, S.H., and Lin, M.R. (2006). Use of the WHOQOL-BREF for evaluating persons with traumatic brain injury. J Neurotrauma 23, 1609–1620.

Claesson, M.J., Jeffery, I.B., Conde, S., Power, S.E., O’Connor, E.M., Cusack, S., Harris, H.M., Coakley, M., Lakshminarayanan, B., O’Sullivan, O., et al. (2012). Gut microbiota composition correlates with diet and health in the elderly. Nature 488, 178–184.

Cryan, J.F., O’Riordan, K.J., Sandhu, K., Peterson, V., and Dinan, T.G. (2020). The gut microbiome in neurological disorders. Lancet Neurol 19, 179–194.

Dai, M., Liu, Y., Chen, W., Buch, H., Shan, Y., Chang, L., Bai, Y., Shen, C., Zhang, X., Huo, Y., et al. (2019). Rescue fecal microbiota transplantation for antibiotic-associated diarrhea in critically ill patients. Crit Care 23, 324.

de Groot, P., Scheithauer, T., Bakker, G.J., Prodan, A., Levin, E., Khan, M.T., Herrema, H., Ackermans, M., Serlie, M.J.M., de Brauw, M., et al. (2020). Donor metabolic characteristics drive effects of faecal microbiota transplantation on recipient insulin sensitivity, energy expenditure and intestinal transit time. Gut 69, 502–512.

Deng, X., Zhang, C., Wang, P., Wei, W., Shi, X., Wang, P., Yang, J., Wang, L., Tang, S., Fang, Y., et al. (2022). Cardiovascular benefits of empagliflozin are associated with gut microbiota and plasma metabolites in type 2 diabetes. J Clin Endocrinol Metab.

Derrien, M., Turroni, F., Ventura, M., and van Sinderen, D. (2022). Insights into endogenous Bifidobacterium species in the human gut microbiota during adulthood. Trends Microbiol.

Dewanjee, S., Das, S., Das, A.K., Bhattacharjee, N., Dihingia, A., Dua, T.K., Kalita, J., and Manna, P. (2018). Molecular mechanism of diabetic neuropathy and its pharmacotherapeutic targets. Eur J Pharmacol 833, 472–523.

Ding, X., Li, Q., Li, P., Zhang, T., Cui, B., Ji, G., Lu, X., and Zhang, F. (2019). Long-Term Safety and Efficacy of Fecal Microbiota Transplant in Active Ulcerative Colitis. Drug Saf 42, 869–880.

Dyck, P.J., Davies, J.L., Litchy, W.J., and O’Brien, P.C. (1997). Longitudinal assessment of diabetic polyneuropathy using a composite score in the Rochester Diabetic Neuropathy Study cohort. Neurology 49, 229–239.

Fan, B., Li, C., Szalad, A., Wang, L., Pan, W., Zhang, R., Chopp, M., Zhang, Z.G., and Liu, X.S. (2020). Mesenchymal stromal cell-derived exosomes ameliorate peripheral neuropathy in a mouse model of diabetes. Diabetologia 63, 431–443.

Fang, Y., Zhang, C., Shi, H., Wei, W., Shang, J., Zheng, R., Yu, L., Wang, P., Yang, J., Deng, X., et al. (2021). Characteristics of the Gut Microbiota and Metabolism in Patients With Latent Autoimmune Diabetes in Adults: A Case-Control Study. Diabetes Care 44, 2738–2746.

Feldman, E.L., Nave, K.A., Jensen, T.S., and Bennett, D.L.H. (2017). New Horizons in Diabetic Neuropathy: Mechanisms, Bioenergetics, and Pain. Neuron 93, 1296–1313.

Forster, S.C., Kumar, N., Anonye, B.O., Almeida, A., Viciani, E., Stares, M.D., Dunn, M., Mkandawire, T.T., Zhu, A., Shao, Y., et al. (2019). A human gut bacterial genome and culture collection for improved metagenomic analyses. Nat Biotechnol 37, 186–192.

Fung, T.C., Olson, C.A., and Hsiao, E.Y. (2017). Interactions between the microbiota, immune and nervous systems in health and disease. Nat Neurosci 20, 145–155.

Gomes, A.C., Hoffmann, C., and Mota, J.F. (2018). The human gut microbiota: Metabolism and perspective in obesity. Gut Microbes 9, 308–325.

Goss, J.R., Goins, W.F., Lacomis, D., Mata, M., Glorioso, J.C., and Fink, D.J. (2002). Herpes simplex-mediated gene transfer of nerve growth factor protects against peripheral neuropathy in streptozotocin-induced diabetes in the mouse. Diabetes 51, 2227–2232.

Gylfadottir, S.S., Christensen, D.H., Nicolaisen, S.K., Andersen, H., Callaghan, B.C., Itani, M., Khan, K.S., Kristensen, A.G., Nielsen, J.S., Sindrup, S.H., et al. (2020). Diabetic polyneuropathy and pain, prevalence, and patient characteristics: a cross-sectional questionnaire study of 5,514 patients with recently diagnosed type 2 diabetes. Pain 161, 574–583.

Hanssen, N.M.J., de Vos, W.M., and Nieuwdorp, M. (2021). Fecal microbiota transplantation in human metabolic diseases: From a murky past to a bright future? Cell Metab 33, 1098–1110.

Herder, C., Kannenberg, J.M., Huth, C., Carstensen-Kirberg, M., Rathmann, W., Koenig, W., Heier, M., Puttgen, S., Thorand, B., Peters, A., et al. (2017). Proinflammatory Cytokines Predict the Incidence and Progression of Distal Sensorimotor Polyneuropathy: KORA F4/FF4 Study. Diabetes Care 40, 569–576.

Hicks, C.W., and Selvin, E. (2019). Epidemiology of Peripheral Neuropathy and Lower Extremity Disease in Diabetes. Curr Diab Rep 19, 86.

Holvoet, T., Joossens, M., Vazquez-Castellanos, J.F., Christiaens, E., Heyerick, L., Boelens, J., Verhasselt, B., van Vlierberghe, H., De Vos, M., Raes, J., et al. (2021). Fecal Microbiota Transplantation Reduces Symptoms in Some Patients With Irritable Bowel Syndrome With Predominant Abdominal Bloating: Short- and Long-term Results From a Placebo-Controlled Randomized Trial. Gastroenterology 160, 145–157 e148.

Joly, A., Leulier, F., and De Vadder, F. (2021). Microbial Modulation of the Development and Physiology of the Enteric Nervous System. Trends Microbiol 29, 686–699.

Kang, D.D., Li, F., Kirton, E., Thomas, A., Egan, R., An, H., and Wang, Z. (2019). MetaBAT 2: an adaptive binning algorithm for robust and efficient genome reconstruction from metagenome assemblies. PeerJ 7, e7359.

Kim, J.E., Kim, H.E., Cho, H., Park, J.I., Kwak, M.J., Kim, B.Y., Yang, S.H., Lee, J.P., Kim, D.K., Joo, K.W., et al. (2020). Effect of the similarity of gut microbiota composition between donor and recipient on graft function after living donor kidney transplantation. Sci Rep 10, 18881.

Klindworth, A., Pruesse, E., Schweer, T., Peplies, J., Quast, C., Horn, M., and Glockner, F.O. (2013). Evaluation of general 16S ribosomal RNA gene PCR primers for classical and next-generation sequencing-based diversity studies. Nucleic Acids Res 41, e1.

Koh, A., De Vadder, F., Kovatcheva-Datchary, P., and Backhed, F. (2016). From Dietary Fiber to Host Physiology: Short-Chain Fatty Acids as Key Bacterial Metabolites. Cell 165, 1332–1345.

Langmead, B., and Salzberg, S.L. (2012). Fast gapped-read alignment with Bowtie 2. Nat Methods 9, 357–359.

Li, J., Zhang, H., Xie, M., Yan, L., Chen, J., and Wang, H. (2013). NSE, a potential biomarker, is closely connected to diabetic peripheral neuropathy. Diabetes Care 36, 3405–3410.

Li, Y., Lv, L., Ye, J., Fang, D., Shi, D., Wu, W., Wang, Q., Wu, J., Yang, L., Bian, X., et al. (2019). Bifidobacterium adolescentis CGMCC 15058 alleviates liver injury, enhances the intestinal barrier and modifies the gut microbiota in D-galactosamine-treated rats. Appl Microbiol Biotechnol 103, 375–393.

Liu, F., Li, J., Feng, G., and Li, Z. (2016). New Genomic Insights into “Entotheonella” Symbionts in Theonella swinhoei: Mixotrophy, Anaerobic Adaptation, Resilience, and Interaction. Front Microbiol 7, 1333.

Long, C., Yu, Y., Cui, B., Jagessar, S.A.R., Zhang, J., Ji, G., Huang, G., and Zhang, F. (2018). A novel quick transendoscopic enteral tubing in mid-gut: technique and training with video. BMC Gastroenterol 18, 37.

Lv, S.L., Fang, C., Hu, J., Huang, Y., Yang, B., Zou, R., Wang, F.Y., and Zhao, H.Q. (2015). Assessment of Peripheral Neuropathy Using Measurement of the Current Perception Threshold with the Neurometer(R) in patients with type 1 diabetes mellitus. Diabetes Res Clin Pract 109, 130–134.

Maioli, T.U., Borras-Nogues, E., Torres, L., Barbosa, S.C., Martins, V.D., Langella, P., Azevedo, V.A., and Chatel, J.M. (2021). Possible Benefits of Faecalibacterium prausnitzii for Obesity-Associated Gut Disorders. Front Pharmacol 12, 740636.

Mallick, H., Rahnavard, A., McIver, L.J., Ma, S., Zhang, Y., Nguyen, L.H., Tickle, T.L., Weingart, G., Ren, B., Schwager, E.H., et al. (2021). Multivariable association discovery in population-scale meta-omics studies. PLoS Comput Biol 17, e1009442.

McGregor, C.E., and English, A.W. (2018). The Role of BDNF in Peripheral Nerve Regeneration: Activity-Dependent Treatments and Val66Met. Front Cell Neurosci 12, 522.

Metwaly, A., Reitmeier, S., and Haller, D. (2022). Microbiome risk profiles as biomarkers for inflammatory and metabolic disorders. Nat Rev Gastroenterol Hepatol 19, 383–397.

Morais, L.H., Schreiber, H.L., and Mazmanian, S.K. (2020). The gut microbiota-brain axis in behaviour and brain disorders. Nat Rev Microbiol.

Olesen, S.W., and Gerardin, Y. (2021). Re-Evaluating the Evidence for Faecal Microbiota Transplantation ‘Super-Donors’ in Inflammatory Bowel Disease. J Crohns Colitis 15, 453–461.

Olm, M.R., Brown, C.T., Brooks, B., and Banfield, J.F. (2017). dRep: a tool for fast and accurate genomic comparisons that enables improved genome recovery from metagenomes through de-replication. Isme j 11, 2864–2868.

Pan, F., Zhang, L., Li, M., Hu, Y., Zeng, B., Yuan, H., Zhao, L., and Zhang, C. (2018). Predominant gut Lactobacillus murinus strain mediates anti-inflammaging effects in calorie-restricted mice. Microbiome 6, 54.

Pane, K., Boccella, S., Guida, F., Franzese, M., Maione, S., and Salvatore, M. (2022). Role of gut microbiota in neuropathy and neuropathic pain states: A systematic preclinical review. Neurobiol Dis 170, 105773.

Parks, D.H., Imelfort, M., Skennerton, C.T., Hugenholtz, P., and Tyson, G.W. (2015). CheckM: assessing the quality of microbial genomes recovered from isolates, single cells, and metagenomes. Genome Res 25, 1043–1055.

Peng, Y., Leung, H.C., Yiu, S.M., and Chin, F.Y. (2012). IDBA-UD: a de novo assembler for single-cell and metagenomic sequencing data with highly uneven depth. Bioinformatics 28, 1420–1428.

Petersen, E.A., Stauss, T.G., Scowcroft, J.A., Brooks, E.S., White, J.L., Sills, S.M., Amirdelfan, K., Guirguis, M.N., Xu, J., Yu, C., et al. (2021). Effect of High-frequency (10-kHz) Spinal Cord Stimulation in Patients With Painful Diabetic Neuropathy: A Randomized Clinical Trial. JAMA Neurol 78, 687–698.

Pop-Busui, R., Boulton, A.J., Feldman, E.L., Bril, V., Freeman, R., Malik, R.A., Sosenko, J.M., and Ziegler, D. (2017). Diabetic Neuropathy: A Position Statement by the American Diabetes Association. Diabetes Care 40, 136–154.

Quast, C., Pruesse, E., Yilmaz, P., Gerken, J., Schweer, T., Yarza, P., Peplies, J., and Glockner, F.O. (2013). The SILVA ribosomal RNA gene database project: improved data processing and web-based tools. Nucleic Acids Res 41, D590–596.

Sanna, S., van Zuydam, N.R., Mahajan, A., Kurilshikov, A., Vich Vila, A., Võsa, U., Mujagic, Z., Masclee, A.A.M., Jonkers, D., Oosting, M., et al. (2019). Causal relationships among the gut microbiome, short-chain fatty acids and metabolic diseases. Nat Genet 51, 600–605.

Saxena, S., Carlson, D., Billington, R., and Life, W.G.W.H.O.Q.O. (2001). The WHO quality of life assessment instrument (WHOQOL-Bref): the importance of its items for cross-cultural research. Qual Life Res 10, 711–721.

Schlesinger, S., Herder, C., Kannenberg, J.M., Huth, C., Carstensen-Kirberg, M., Rathmann, W., Bonhof, G.J., Koenig, W., Heier, M., Peters, A., et al. (2019). General and Abdominal Obesity and Incident Distal Sensorimotor Polyneuropathy: Insights Into Inflammatory Biomarkers as Potential Mediators in the KORA F4/FF4 Cohort. Diabetes Care 42, 240–247.

Seemann, T. (2014). Prokka: rapid prokaryotic genome annotation. Bioinformatics 30, 2068–2069.

Shi, Y., Zhang, L., Do, K.A., Peterson, C.B., and Jenq, R. (2020). aPCoA: Covariate Adjusted Principal Coordinates Analysis. Bioinformatics.

Slangen, R., Schaper, N.C., Faber, C.G., Joosten, E.A., Dirksen, C.D., van Dongen, R.T., Kessels, A.G., and van Kleef, M. (2014). Spinal cord stimulation and pain relief in painful diabetic peripheral neuropathy: a prospective two-center randomized controlled trial. Diabetes Care 37, 3016–3024.

Sloan, G., Selvarajah, D., and Tesfaye, S. (2021). Pathogenesis, diagnosis and clinical management of diabetic sensorimotor peripheral neuropathy. Nat Rev Endocrinol 17, 400–420.

Sorboni, S.G., Moghaddam, H.S., Jafarzadeh-Esfehani, R., and Soleimanpour, S. (2022). A Comprehensive Review on the Role of the Gut Microbiome in Human Neurological Disorders. Clin Microbiol Rev 35, e0033820.

Tang, S.S., Liang, C.H., Liu, Y.L., Wei, W., Deng, X.R., Shi, X.Y., Wang, L.M., Zhang, L.J., and Yuan, H.J. (2022). Intermittent hypoxia is involved in gut microbial dysbiosis in type 2 diabetes mellitus and obstructive sleep apnea-hypopnea syndrome. World journal of gastroenterology 28, 2320–2333.

Tansey, M.G., Wallings, R.L., Houser, M.C., Herrick, M.K., Keating, C.E., and Joers, V. (2022). Inflammation and immune dysfunction in Parkinson disease. Nat Rev Immunol.

Thevaranjan, N., Puchta, A., Schulz, C., Naidoo, A., Szamosi, J.C., Verschoor, C.P., Loukov, D., Schenck, L.P., Jury, J., Foley, K.P., et al. (2017). Age-Associated Microbial Dysbiosis Promotes Intestinal Permeability, Systemic Inflammation, and Macrophage Dysfunction. Cell Host Microbe 21, 455–466 e454.

Tilg, H., Zmora, N., Adolph, T.E., and Elinav, E. (2020). The intestinal microbiota fuelling metabolic inflammation. Nat Rev Immunol 20, 40–54.

Tirosh, A., Calay, E.S., Tuncman, G., Claiborn, K.C., Inouye, K.E., Eguchi, K., Alcala, M., Rathaus, M., Hollander, K.S., Ron, I., et al. (2019). The short-chain fatty acid propionate increases glucagon and FABP4 production, impairing insulin action in mice and humans. Sci Transl Med 11.

Vincent, A.M., Calabek, B., Roberts, L., and Feldman, E.L. (2013). Biology of diabetic neuropathy. Handb Clin Neurol 115, 591–606.

Vital, M., Howe, A.C., and Tiedje, J.M. (2014). Revealing the bacterial butyrate synthesis pathways by analyzing (meta)genomic data. mBio 5, e00889.

Wang, Y., Ye, X., Ding, D., and Lu, Y. (2020). Characteristics of the intestinal flora in patients with peripheral neuropathy associated with type 2 diabetes. J Int Med Res 48, 300060520936806.

Waters, J.L., and Ley, R.E. (2019). The human gut bacteria Christensenellaceae are widespread, heritable, and associated with health. BMC Biol 17, 83.

Wu, G., Xu, T., Zhao, N., Lam, Y.Y., Ding, X., Wei, D., Fan, J., Shi, Y., Li, X., Li, M., et al. (2022). Two Competing Guilds as a Core Microbiome Signature for Chronic Diseases. bioRxiv, 2022.2005.2002.490290.

Wu, G., Zhao, N., Zhang, C., Lam, Y.Y., and Zhao, L. (2021). Guild-based analysis for understanding gut microbiome in human health and diseases. Genome Med 13, 22.

Xiao, S., Jiang, S., Qian, D., and Duan, J. (2020). Modulation of microbially derived short-chain fatty acids on intestinal homeostasis, metabolism, and neuropsychiatric disorder. Appl Microbiol Biotechnol 104, 589–601.

Zhang, C., and Zhao, L. (2016). Strain-level dissection of the contribution of the gut microbiome to human metabolic disease. Genome Med 8, 41.

Zhang, C.H., Lv, X., Du, W., Cheng, M.J., Liu, Y.P., Zhu, L., and Hao, J. (2019). The Akt/mTOR cascade mediates high glucose-induced reductions in BDNF via DNMT1 in Schwann cells in diabetic peripheral neuropathy. Exp Cell Res 383, 111502.

Zhang, L., Liu, C., Jiang, Q., and Yin, Y. (2021). Butyrate in Energy Metabolism: There Is Still More to Learn. Trends Endocrinol Metab 32, 159–169.

Zhang, T., Lu, G., Zhao, Z., Liu, Y., Shen, Q., Li, P., Chen, Y., Yin, H., Wang, H., Marcella, C., et al. (2020). Washed microbiota transplantation vs. manual fecal microbiota transplantation: clinical findings, animal studies and in vitro screening. Protein Cell 11, 251–266.

Zhao, C.G., Sun, W., Ju, F., Jiang, S., Wang, H., Sun, X.L., Mou, X., and Yuan, H. (2021). Analgesic Effects of Navigated Repetitive Transcranial Magnetic Stimulation in Patients With Acute Central Poststroke Pain. Pain Ther 10, 1085–1100.

Zhao, L., Zhang, F., Ding, X., Wu, G., Lam, Y.Y., Wang, X., Fu, H., Xue, X., Lu, C., Ma, J., et al. (2018). Gut bacteria selectively promoted by dietary fibers alleviate type 2 diabetes. Science 359, 1151–1156.

