## Supplementary Figures 1-8 and Tables 1-4 for "Gut microbiota modulates distal symmetric polyneuropathy in diabetic patients"

**Figure S1**

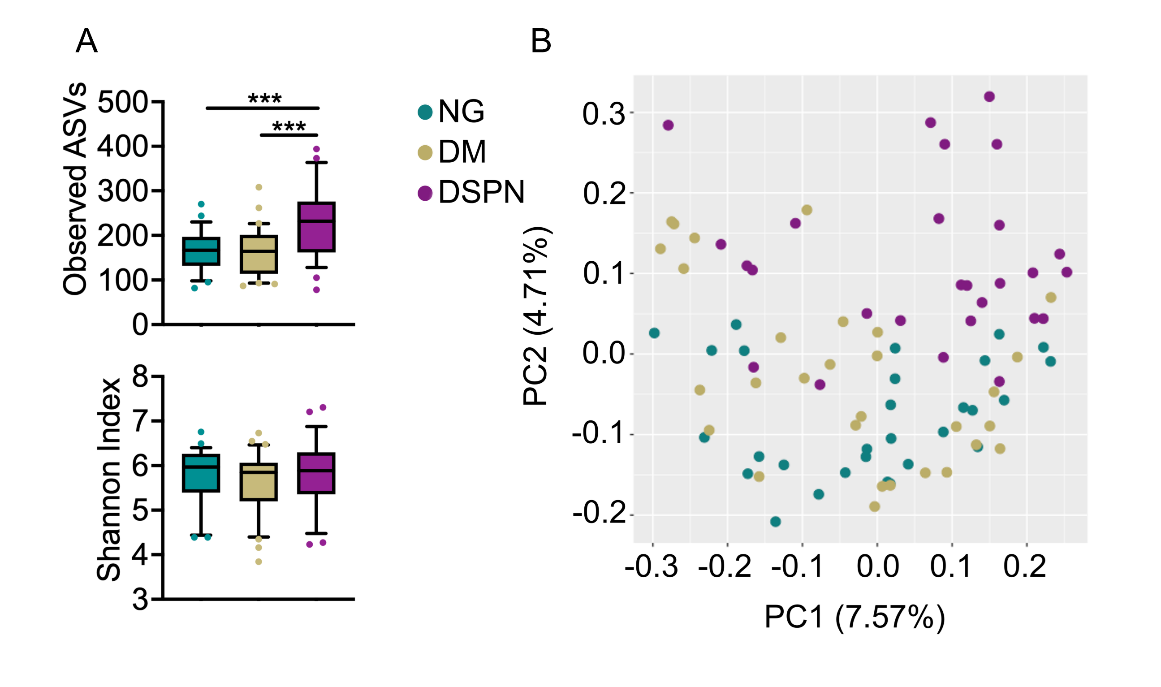

**Figure S1.** **Gut microbiota showed significant differences among NG, DM and DSPN cohorts. (A)** Richness (Observed ASVs) and diversity (Shannon index) of the gut microbiota. The line in the middle of the box is plotted at the median, the inferior and superior limits of the box correspond to the 25th and 75th percentiles, the whiskers correspond to the 10^th^ and 90^th^ percentiles, and outliers are denoted. Kruskal-Wallis test followed by Dunn’s post hoc was used to compare the three groups. *** *P* < 0.001**. (B)** Principal coordinate analysis (PCoA) of Jaccard distances at the ASV-level. NG indicates the subjects with normal glucose level (n = 29), DM indicates type 2 diabetes patients without peripheral neuropathy group (n = 30), DSPN indicates type 2 diabetes patients with peripheral neuropathy group (n = 27).

**Figure S2**

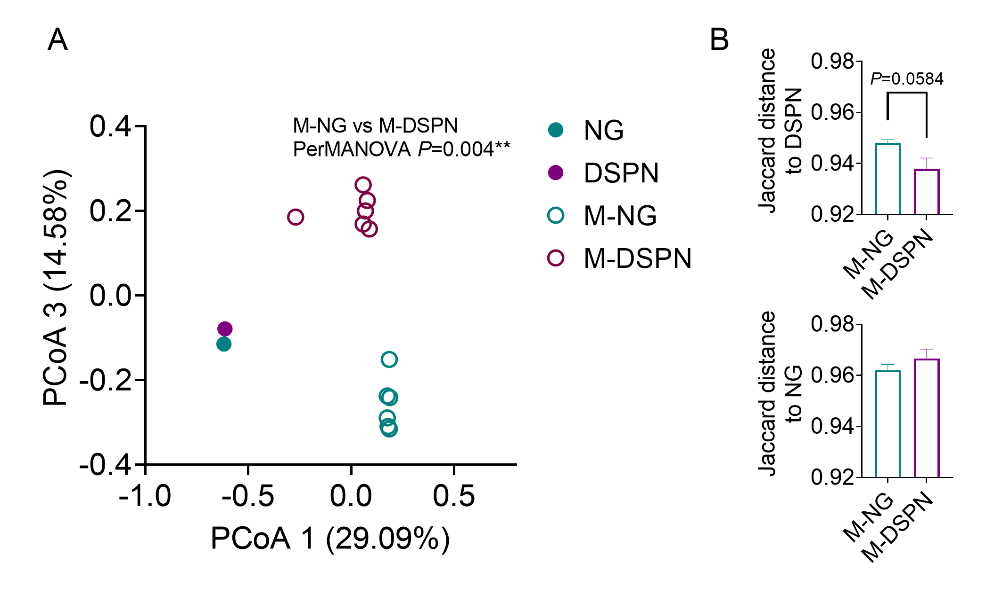

**Figure S2. The changes in the gut microbiota of db/db mice subjected to FMT.**

The structure changes in the gut microbiota in db/db mice subjected to FMT. **(A)** Principal coordinate analysis plot and PerMANOVA test result based on Jaccard distance of gut microbiota; **(B)** Jaccard distance between the mouse and donor in each group. Data are presented as the mean ± s.e.m, the Jaccard distance was tested by Mann-Whitney U test. ** *P* < 0.01. NG indicates normal glucose level donor (n = 1), DSPN indicates DM with peripheral neuropathy donor (n = 1), M-NG indicates db/db mice received microbiota from NG donor (n = 6), M-DSPN indicates db/db mice received microbiota from DSPN donor (n = 6).

**Figure S3**

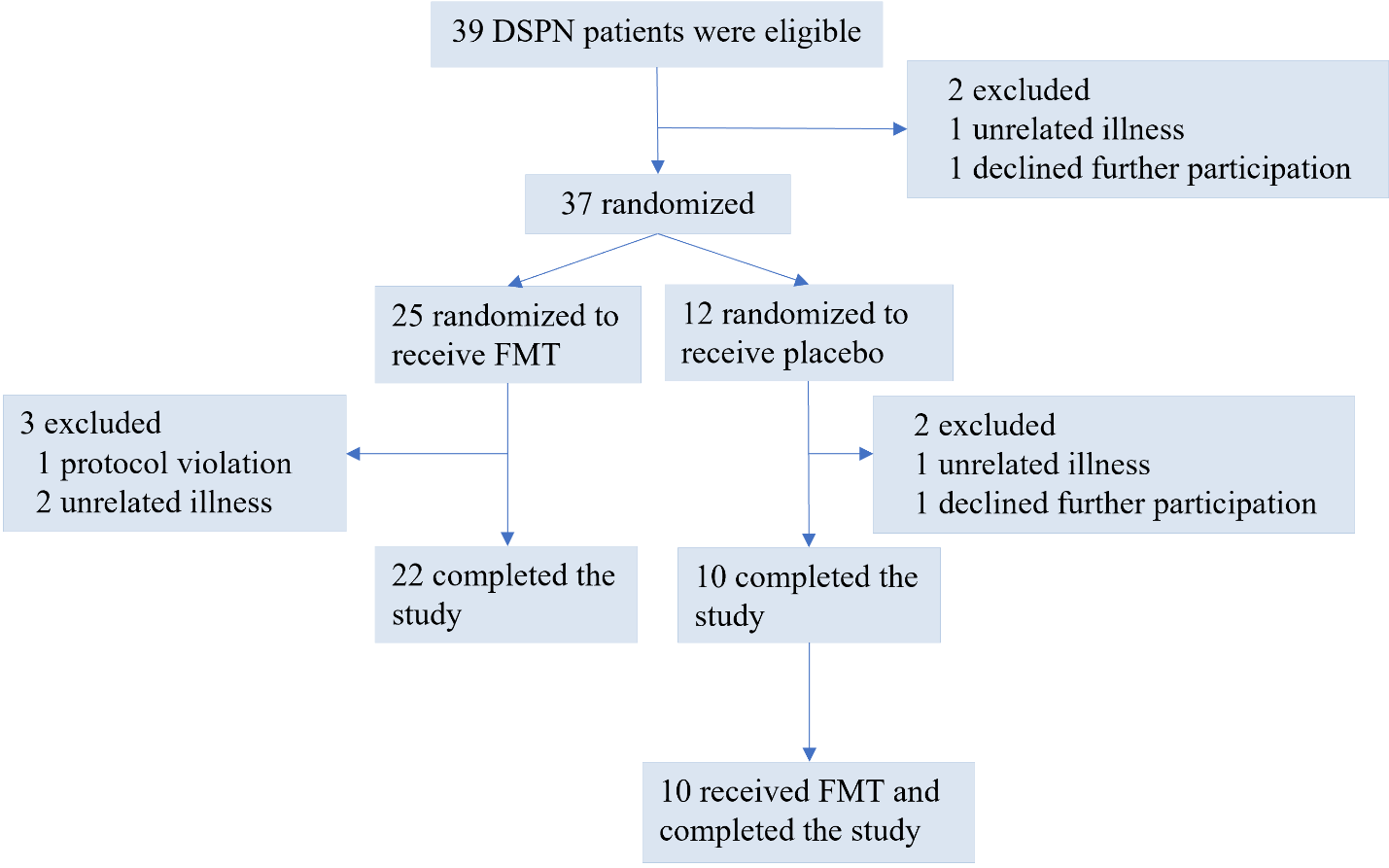

**Figure S3. Participant Flow of the RCT.** FMT: fecal microbiota transplantation.

**Figure S4**

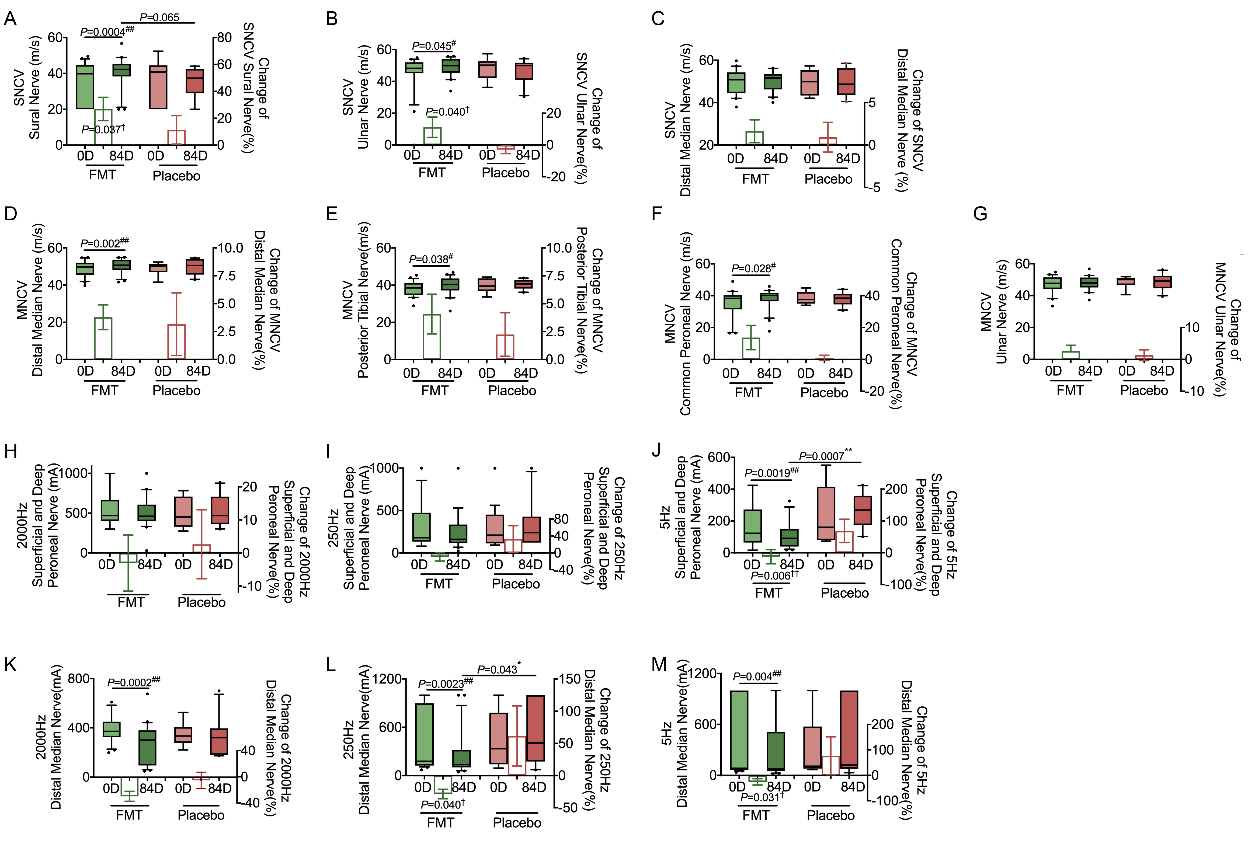

**Figure S4.** **Alleviation of NCV and CPT in DSPN patients subjected to FMT. (A)** SNCV of sural nerve; **(B)** SNCV of the ulnar nerve; **(C)** SNCV of the distal median nerve; **(D)** MNCV of the distal median nerve; **(E)** MNCV of the posterior tibial nerve; **(F)** MNCV of the common peroneal nerve; **(G)** MNCV of the ulnar nerve; **(H)** CPT level of superficial and deep peroneal nerve measured at 2000 Hz; **(I)** CPT level of superficial and deep peroneal nerve measured at 250 Hz; **(J)** CPT level of superficial and deep peroneal nerve measured at 5 Hz; **(K)** CPT level of distal median nerve measured at 2000 Hz; **(L)** CPT level of distal median nerve measured at 250 Hz; **(M)** CPT level of distal median nerve measured at 5 Hz. 0D indicates baseline, and 84D indicates 84 days after FMT. The bars represent the mean change from the baseline value per group, with the corresponding s.e.m. Mann-Whitney U test was used to analyze differences in the changes between the FMT and placebo groups (intergroup changes). ^†^ *P* <0.05 and ^††^ *P* <0.01. In the box plots, the line in the middle of the box is plotted at the median, and the inferior and superior limits of the box correspond to the 25th and 75th percentiles, respectively. The whiskers correspond to the 10th and 90th percentiles, and outliers are denoted. Mann-Whitney U test was used to analyze differences between the two groups. * *P* < 0.05. **Wilcoxon matched-pairs signed rank test** was used to analyze each pairwise comparison within each group. ^#^ *P* < 0.05 and ^##^ *P* < 0.01. FMT group, n = 22; placebo group, n = 10. FMT: fecal microbiota transplantation; SNCV: sensory nerve conduction velocity; MNCV: motor nerve conduction velocity; CPT: current perception threshold.

**Figure S5**

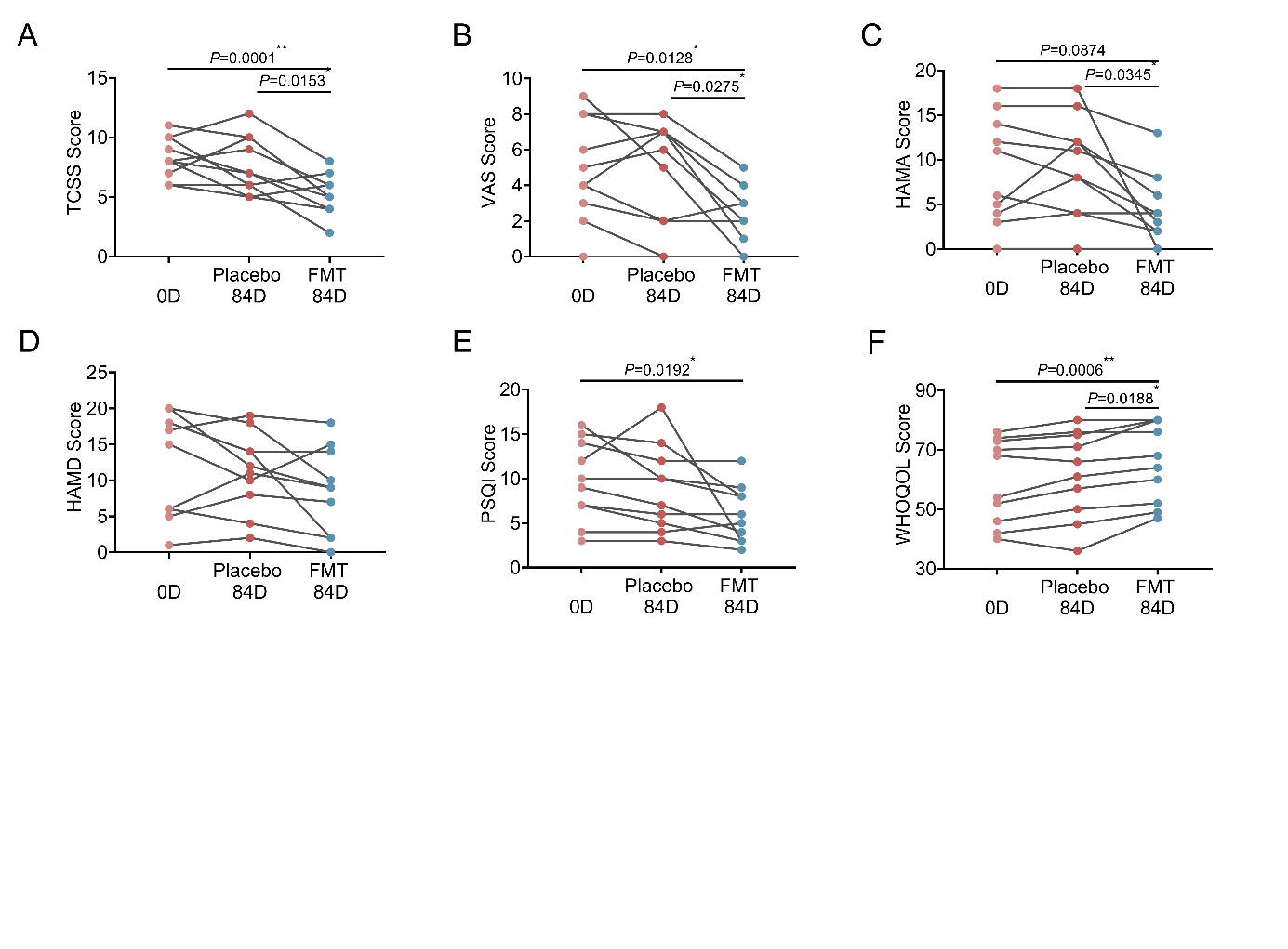

**Figure S5.** **Changes of the severity of peripheral neuropathy, anxiety, depression, sleep and life quality in Placebo group during the RCT and the post-RCT study of transplantation with transplants from healthy donors. (A)** Toronto clinical scoring system **(**TCSS) score. **(B)** Visual analogue scale (VAS) score. **(C)** Hamilton anxiety scale (HAMA) score. **(D)** Hamilton depression rating scale (HAMD) score. (**E**) Pittsburgh sleep quality index (PSQI) score. (**F**) World health organization’s quality of life (WHOQOL)-BREF score. One-Way RM ANOVA test was used to analyze differences between the three time points. * *P* < 0.05, ** *P* < 0.01. 0D indicates baseline of RCT (n = 10), Placebo 84D indicates 84 days after transplantation with placebo in the RCT study (n = 10) and FMT 84D indicates 84 days after transplantation with transplants from healthy donors in the post-RCT study (n = 10).

**Figure S6**

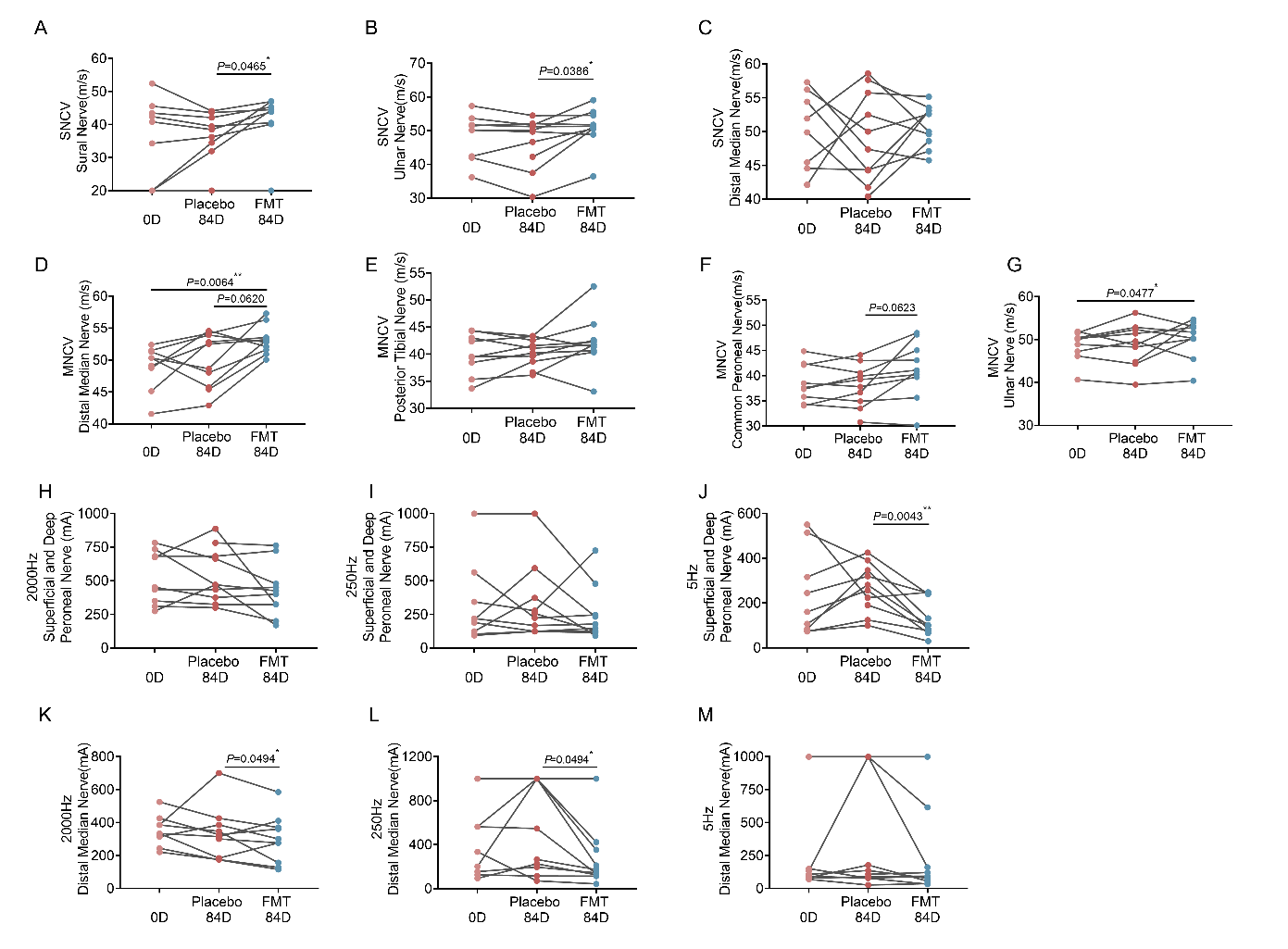

**Figure S6.** **Changes of NCV and CPT of patients in Placebo group during the RCT and the post-RCT study of transplantation with transplants from healthy donors.** (**A**) SNCV of sural nerve. (**B**) SNCV of the ulnar nerve. (**C**) SNCV of the distal median nerve. (**D**) MNCV of the distal median nerve. (**E**) MNCV of the posterior tibial nerve; **(F)** MNCV of the common peroneal nerve; **(G)** MNCV of the ulnar nerve; **(H)** CPT level of superficial and deep peroneal nerve measured at 2000 Hz; **(I)** CPT level of superficial and deep peroneal nerve measured at 250 Hz; **(J)** CPT level of superficial and deep peroneal nerve measured at 5 Hz; **(K)** CPT level of distal median nerve measured at 2000 Hz; **(L)** CPT level of distal median nerve measured at 250 Hz; **(M)** CPT level of distal median nerve measured at 5 Hz. One-Way RM ANOVA test was used to analyze differences between the three time points. * *P* < 0.05, ** *P* < 0.01. 0D indicates baseline of RCT (n = 10), Placebo 84D indicates 84 days after transplantation with placebo in the RCT study (n = 10) and FMT 84D indicates 84 days after transplantation with transplants from healthy donors in the post-RCT study (n = 10). FMT: fecal microbiota transplantation; SNCV: sensory nerve conduction velocity; MNCV: motor nerve conduction velocity; CPT: current perception threshold.

**Figure S7**

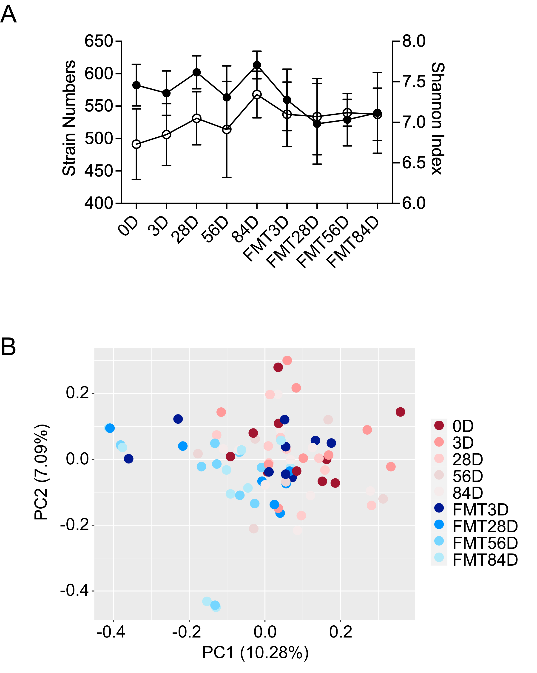

**Figure S7. The gut microbiota population of the placebo group patients was unchanged during the RCT study and significantly shifted in the post-RCT study when transplanted with transplants from healthy donors. (A)** Richness (number of strains) and diversity (Shannon index) of the gut microbiota. Data are presented as the mean ± s.e.m. **(B)** Subject adjusted principal coordinate analysis of Jaccard distances at the strain level. 0D indicates baseline (n = 10). 3D (n = 9), 28D (n = 10), 56D (n = 6) and 84D (n = 10) indicates 3 days, 28 days, 56 days and 84 days after transplantation with placebo in RCT study. FMT3D (n = 9), FMT28D (n = 7), FMT56D (n = 9), and FMT84D (n = 10) indicates 3 days, 28 days, 56 days and 84 days after transplantation with transplants from healthy donors in the post-RCT study, respectively.

**Figure S8**

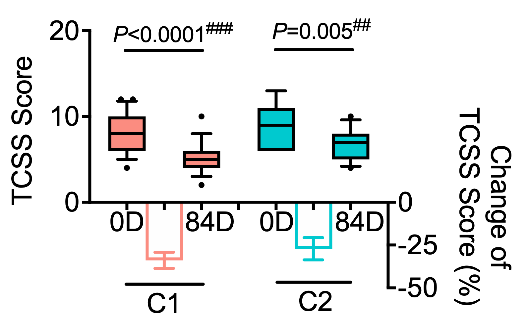

**Figure S8. The enterotypes of transplants did not affect the efficacy of TCSS improvement.** The samples from FMT group in RCT study and samples from Placebo group in the post-RCT study of transplantation with transplants from healthy donors were analyzed together. 0D indicates before transplantation and 84D indicate 84 days after transplantation with transplants from healthy donors. C1 indicates the patients received the transplants belonged to enterotype C1 (n = 21). C2 indicates the patients received the transplants belonged to enterotype C2 (n = 11). In the box plots, the line in the middle of the box is plotted at the median, and the inferior and superior limits of the box correspond to the 25^th^ and 75^th^ percentiles, respectively. The whiskers correspond to the 10^th^ and 90^th^ percentiles, and outliers are denoted. Mann-Whitney U test was used to analyze differences between C1 and C2 groups. Wilcoxon matched-pairs signed rank test was used to analyze each pairwise comparison 0D and 84D within each group. ^##^ *P* < 0.01 and ^###^ *P* < 0.001. The bars represent the mean change from the baseline value per group, with the corresponding s.e.m. Mann-Whitney U test was used to analyze differences in the changes between the C1 and C2 groups (intergroup changes).

**Supplementary Table 1.** Clinical characteristics of DM patients with DSPN as compared with DM patients without DSPN and subjects with normal glucose level

| Clinical characteristics | NG | DM | DSPN | *P* Value |
| --- | --- | --- | --- | --- |
| Age (years) | 34.00(28.5-52.5) | 49.97±9.10 | 49.48±10.60^a^ | 0.005 |
| Gender (Females/Males) | 11/18 | 11/19 | 4/23 | 0.108 |
| Duration of diabetes (years) | / | 1.00(0.00-3.50) | 8.93±6.16^d^ | / |
| TCSS | / | 1.00(0.00-2.00） | 9.00±2.48^d^ | / |
| HbA1c (%) | 5.33±0.28 | 8.91±1.87 | 8.14±2.04^c^ | ＜0.001 |
| FBG (mmol/L) | 4.87±0.50 | 7.34±1.58 | 6.37(5.15-9.38)^b^ | ＜0.001 |

Values are expressed as the mean ± s.e.m., median with interquartile range (IQR), or number. One-way ANOVA or Kolmogorov-Smirnov test was used to detect the differences among the three groups. ^a^ *P* < 0.05 vs NG, ^b^ *P* < 0.01 vs NG, ^c^ *P* < 0.001 vs NG. Mann-Whitney U test was used to detect the differences among the two groups. ^d^ *P* < 0.001 vs DM. NG indicates normal glucose level group (n = 29), DM indicates DM patients without peripheral neuropathy group (n = 30), DSPN indicates DM patients with distal symmetric polyneuropathy group (n = 27). TCSS: Toronto Clinical Scoring System; HbA1c, glycated haemoglobin; FBG, fasting blood glucose.

**Supplementary Table 2.** The symptoms and conventional treatments before FMT of the patients who completed the study

| Case | DSPN symptoms | Duration of  DSPN symptoms | Conventional Treatments | | |
| --- | --- | --- | --- | --- | --- |
|  |  |  | Lifestyle modification | Glucose control | Drug intervention* |
| 1 | Pain | 4 months | Yes | Insulin | Duloxetine, Flupentixol and Melitracen, Gabapentin |
| 3 | Numbness +  Pain | 8 years | Yes | Oral hypoglycemic drugs + Insulin | α lipoic acid, Duloxetine, Gabapentin, Mecobalamine |
| 4 | Numbness +  Pain | 9 months | Yes | Oral hypoglycemic drugs | α lipoic acid, Duloxetine, Gabapentin, Pregabalin |
| 5 | Pain | 1 year | Yes | Oral hypoglycemic drugs + Insulin | Duloxetine, Flupentixol and Melitracen, Mecobalamine, Pregabalin |
| 6 | Numbness + Pain | 1 year | Yes | Oral hypoglycemic drugs | α lipoic acid, Duloxetine, Gabapentin, Mecobalamine |
| 8 | Numbness | 1 year | Yes | Oral hypoglycemic drugs + Insulin | α lipoic acid, Mecobalamine |
| 9 | Numbness | 4 years | Yes | Oral hypoglycemic drugs | α lipoic acid, Epalrestat |
| 10 | Numbness +  Pain | 10 years | Yes | Oral hypoglycemic drugs + Insulin | Gabapentin, Mecobalamine, Pregabalin, Venlafaxine |
| 11 | Numbness +  Pain | 1 year | Yes | Insulin | α lipoic acid, Duloxetine, Gabapentin, Mecobalamine |
| 12 | Pain | 2 years | Yes | Insulin | Duloxetine, Flupentixol and Melitracen, Gabapentin, Mecobalamine, Pregabalin |
| 14 | Numbness +  Paraesthesia | 1 year | Yes | Oral hypoglycemic drugs + Insulin | α lipoic acid, Mecobalamine, Pancreatic Kininogenase enteric-coated Tablets |
| 15 | Pain | 2 years | Yes | Oral hypoglycemic drugs | Duloxetine, Gabapentin |
| 16 | Numbness +  Pain | 9 years | Yes | Oral hypoglycemic drugs | α lipoic acid, Epalrestat, Gabapentin, Mecobalamine |
| 17 | Numbness | 3 years | Yes | Oral hypoglycemic drugs + Insulin | α lipoic acid, Epalrestat, Mecobalamine |
| 18 | Numbness +  Pain | 5 years | Yes | Oral hypoglycemic drugs + Insulin | α lipoic acid, Mecobalamine |
| 19 | Numbness | 1 year | Yes | Insulin | α lipoic acid, Pancreatic Kininogenase enteric-coated Tablets |
| 20 | Numbness + Pain | 5 months | Yes | Oral hypoglycemic drugs + Insulin | Epalrestat, Flupentixol and Melitracen, Gabapentin, Venlafaxine |
| 21 | Pain | 9 years | Yes | Oral hypoglycemic drugs+Insulin | Vitamin B6, Gabapentin, Duloxetine, Epalrestat |
| 22 | Numbness + Pain | 6 months | Yes | Oral hypoglycemic drugs+Insulin | α lipoic acid, Mecobalamine, sertraline |
| 23 | Numbness + Pain | 1 year | Yes | Oral hypoglycemic drugs+Insulin | Duloxetine, Gabapentin, Mecobalamine, Epalrestat, Carbamazepine |
| 24 | Numbness + Pain | 4 years | Yes | Oral hypoglycemic drugs+Insulin | α lipoic acid, Mecobalamine, Epalrestat, Oxycodone and Acetaminophen Tablets, Pregabalin, Venlafaxine |
| 25 | Pain | 2 years | Yes | Oral hypoglycemic drugs+Insulin | α lipoic acid, Mecobalamine, sufentanil, Gabapentin |
| 26 | Numbness + Pain | 8 years | Yes | Oral hypoglycemic drugs+Insulin | α lipoic acid, Mecobalamine, Pancreatic Kininogenase enteric-coated Tablets |
| 29 | Numbness | 2 years | Yes | Insulin | Vitamin B6, Mecobalamine, Epalrestat |
| 30 | Numbness + Pain | 5 years | Yes | Insulin | Mecobalamine, Gabapentin, Pancreatic Kininogenase enteric-coated Tablets, Amitriptyline |
| 31 | Numbness | 2 years | Yes | Insulin | α lipoic acid, Epalrestat, Pancreatic Kininogenase enteric-coated Tablets |
| 32 | Numbness | 2 years | Yes | Oral hypoglycemic drugs+Insulin | α lipoic acid, Mecobalamine |
| 33 | Numbness + Pain | 4 months | Yes | Oral hypoglycemic drugs+Insulin | Carbamazepine, Gabapentin, Mecobalamine |
| 34 | Numbness + Pain | 6 months | Yes | Oral hypoglycemic drugs | Gabapentin, α lipoic acid, Mecobalamine, Amitriptyline |
| 35 | Numbness + Pain | 1 year | Yes | Insulin | α lipoic acid, Mecobalamine, Epalrestat, Gabapentin |
| 36 | Numbness + Pain | 5 months | Yes | Insulin | α lipoic acid, Mecobalamine, Pancreatic Kininogenase enteric-coated Tablets, Gabapentin |
| 37 | Numbness + Pain | 8 months | Yes | Oral hypoglycemic drugs+Insulin | Gabapentin, Vitamin B6, Epalrestat |

*According to the guideline of American Diabetes Association (2017) and clinical drug recommendation, at least two drugs were applied more than 3 months. DSPN: distal symmetric polyneuropathy.

**Supplementary Table 3.** Recipient clinical characteristics over time during the RCT study

| Clinical characteristics | FMT | | | Placebo | | | *P* Value | |
| --- | --- | --- | --- | --- | --- | --- | --- | --- |
|  | 0D | 84D | *P* Value | 0D | 84D | *P* Value | 0D | 84D |
| General Information |  |  |  |  |  |  |  |  |
| Age (years) | 48.50±10.71 | / | / | 47.90±11.27 | / | / | 0.886 | / |
| Gender (Females/Males) | 2/20 | / | / | 3/7 | / | / | 0.137 | / |
| Type of Diabetes (T2D/T1D) | 16/6 | / | / | 8/2 | / | / | 0.665 | / |
| Duration of diabetes (years) | 9.05±6.85 | / | / | 6.67±3.61 | / | / | 0.595 | / |
| Insulin dose (U) | 27.17±11.94 | 29.65±10.71 | 0.123 | 26.36±10.21 | 25.90±12.35 | 0.852 | 0.855 | 0.393 |
| Plasma glucose homeostasis |  |  |  |  |  |  |  |  |
| HbA1c (%) | 8.50±2.29 | 8.23±1.70 | 0.253 | 7.96±1.37 | 8.17±1.87 | 0.893 | 0.421 | 0.931 |
| FBG (mmol/L) | 6.31±1.38 | 6.70(5.95,8.80) | 0.140 | 6.33±1.26 | 6.51±1.85 | 0.639 | 0.959 | 0.521 |
| OGTT_0.5h PBG (mmol/L) | 11.06±2.67 | 11.00(9.35,11.40) | 0.398 | 10.67±3.20 | 11.36±1.26 | 0.724 | 0.741 | 0.499 |
| OGTT_1h PBG (mmol/L) | 14.74±2.53 | 13.81±2.73 | 0.975 | 14.18±3.14 | 14.89±2.89 | 0.971 | 0.614 | 0.420 |
| OGTT_2h PBG (mmol/L) | 17.46±3.63 | 15.94±3.65 | 0.776 | 17.71±4.47 | 17.47±5.91 | 0.63 | 0.874 | 0.480 |
| OGTT_3h PBG (mmol/L) | 15.78±3.79 | 14.62±2.96 | 0.587 | 14.38±6.99 | 13.54±6.50 | 0.084 | 0.584 | 0.689 |
| OGTT_Glucose_AUC | 43.24±7.29 | 40.20±7.09 | 0.790 | 42.24±8.38 | 42.53±12.13 | 0.504 | 0.749 | 0.591 |
| Fasting Blood C-peptide (ng/mL) | 0.84±0.72 | 0.60(0.29,1.29) | 0.191 | 0.73±0.61 | 0.71(0.46,2.23) | 0.345 | 0.646 | 0.417 |
| OGTT_0.5h C-peptide (ng/mL) | 1.04(0.77,2.29) | 1.13(0.64,1.81) | 0.388 | 1.48±1.66 | 2.36±1.75 | 0.187 | 0.517 | 0.216 |
| OGTT_1h C-peptide (ng/mL) | 1.45(0.93,3.63) | 1.95±1.40 | 0.552 | 2.02±2.31 | 3.25±2.59 | 0.211 | 0.566 | 0.170 |
| OGTT_2h C-peptide (ng/mL) | 3.24±2.63 | 2.73±1.50 | 0.366 | 3.67±3.83 | 5.51±3.65 | 0.649 | 0.723 | 0.124 |
| OGTT_3h C-peptide (ng/mL) | 3.37±2.66 | 2.85±1.41 | 0.298 | 2.99±2.36 | 4.28±2.17 | 0.525 | 0.704 | 0.100 |
| OGTT C-peptide AUC | 7.78±6.34 | 6.55±3.84 | 0.304 | 7.56±7.37 | 11.50±7.18 | 0.271 | 0.934 | 0.161 |
| Plasma lipid homeostasis |  |  |  |  |  |  |  |  |
| TC (mmol/L) | 4.71±1.30 | 4.75±1.10 | 0.875 | 4.33±0.99 | 4.27±1.02 | 0.661 | 0.414 | 0.256 |
| Trig (mmol/L) | 1.65±0.98 | 1.26(0.83,1.91) | 0.224 | 1.07(0.91,1.88) | 1.14±0.61 | 0.139 | 0.440 | 0.228 |
| HDL (mmol/L) | 1.12±0.22 | 1.24±0.29 | 0.004** | 1.21±0.46 | 1.29±0.47 | 0.459 | 0.583 | 0.769 |
| LDL (mmol/L) | 2.74±1.03 | 2.76±0.86 | 0.746 | 2.41±0.79 | 2.42±0.7 0 | 0.902 | 0.378 | 0.289 |
| Liver function |  |  |  |  |  |  |  |  |
| ALT (U/L) | 16.50(12.60,26.75) | 18.80(12.45,26.53) | 0.768 | 13.20(10.68,26.60) | 18.85(13.05,21.65) | 0.374 | 0.416 | 0.968 |
| GGT (U/L) | 20.35(14.35,25.63) | 22.35±9.70 | 0.917 | 23.10(16.35,61.08) | 22.15(14.23,47.65) | 0.038 | 0.300 | 0.597 |
| Renal function |  |  |  |  |  |  |  |  |
| Creatinine (umol/L) | 60.00(47.00,85.00) | 57.00(47.50,87.75) | 0.643 | 59.38±17.43 | 57.22±18.66 | 0.639 | 0.864 | 0.459 |
| UA (umol/L) | 278.00(223.50,324.50) | 293.77±75.36 | 0.794 | 246.38±68.39 | 272.56±88.30 | 0.423 | 0.172 | 0.503 |
| Anthropometric markers |  |  |  |  |  |  |  |  |
| BMI (kg/m^2^) | 22.07±3.94 | 22.12±3.32 | 0.844 | 22.87±4.49 | 22.55±4.36 | 0.539 | 0.614 | 0.764 |
| SBP (mmHg) | 121.73±15.68 | 123.68±24.79 | 0.652 | 121.70±31.92 | 114.30±20.76 | 0.331 | 0.998 | 0.307 |
| DBP (mmHg) | 77.18±8.21 | 74.14±14.82 | 0.269 | 81.20±17.07 | 75.40±11.21 | 0.117 | 0.494 | 0.812 |
| Other biomarker |  |  |  |  |  |  |  |  |
| VB12 (pg/mL) | 496.1±150.58 | 516.4±150.94 | 0.487 | 490.5±251.59 | 454.1±135.83 | 0.552 | 0.940 | 0.281 |

Values are expressed as the mean ± s.e.m., median with interquartile range (IQR), or number. Student’s t-test (two-tailed), Wilcoxon matched-pair signed-rank test and Pearson's chi-squared test were used to detect the differences between the FMT and the placebo groups at 0D and 84D. Paired t-test (two-tailed) was used to analyze each pairwise comparison within each group. ** *P* < 0.01. This table includes all participants that completed the RCT study: FMT n = 22, Placebo n = 10. FMT: fecal microbiota transplantation; HbA1c, glycated haemoglobin; FBG, fasting blood glucose; PBG, postprandial blood glucose; TC, total cholesterol; Trig, triglyceride; HDL, high-density lipoprotein; LDL, low-density lipoprotein; ALT, Alanine aminotransferase; GGT, Glutamyl transpeptidase; UA, Uric Acid; BMI, Body Mass Index; SBP, systolic blood; DBP, diastolic blood pressure, VB12, vitamin B12.

**Supplementary Table 4.** Details of all adverse events and relevance to FMT during the RCT study

| **FMT Group (N=22)** | | | | | **Placebo Group (N=10)** | | | | | |
| --- | --- | --- | --- | --- | --- | --- | --- | --- | --- | --- |
| **Pt** | **Time post FMT (day)** | **Adverse events** | **Grade** * | **Causality between adverse events and FMT** | **Pt** | **Time post FMT (day)** | **Adverse events** | | **Grade** * | **Causality between adverse events and FMT** |
| 1 | <7 | Diarrhea | 1 | Probable | 8 | 56 | Diarrhea | 1 | | Possible |
| 3 | 28 | Flu like symptoms | 1 | Possible | 11 | 56 | Vaginal inflammation | 1 | | Possible |
|  | 56 | Flu like symptoms | 1 | Possible | 13 | 28 | Flu like symptoms | 1 | | Possible |
| 4 | 28 | Fever | 1 | Possible |  | 28 | Fever | 2 | | Possible |
| 5 | 56 | Flu like symptoms | 1 | Possible | 15 | 56 | Flu like symptoms | 1 | | Possible |
| 9 | <7 | Diarrhea | 2 | Probable |  |  |  |  | |  |
| 14 | <7 | Generalized edema + Herpes simplex reactivation | 2 | Probable |  |  |  |  | |  |
| 32 | <7 | Diarrhea | 1 | Probable |  |  |  |  | |  |
|  | <7 | Flu like symptoms | 1 | Possible |  |  |  |  | |  |
| 36 | <7 | Diarrhea | 1 | Probable |  |  |  |  | |  |

* Grade according to the Common Terminology Criteria for Adverse Events, version 5.0.

**Supplementary Table 5.** The samples for metagenomic sequencing of RCT and post-RCT (Separate document)

**Supplementary Table 6.** The number of high quality metagenomic reads of 212 samples (Separate document)

**Supplementary Table 7.** The assembly results of 1,572 high quality draft genomes of microorganism from all the samples (Separate document)

**Supplementary Table 8.** The taxonomic annotation information of 1,999 non-redundant genomes (considered as strains) (Separate document)

**Supplementary Table 9.** The 54 genomes correlated with TCSS score (Separate document)
